# Socio-demographic inequalities in COVID-19 health care access and experiences in the United Kingdom: intersectional and mixed-methods analyses of open and closed questions in a prospective cohort study

**DOI:** 10.1101/2025.05.29.25328562

**Authors:** Nathan J. Cheetham, Anoushka Beattie, Alastair B. Comery, Vicky Bowyer, The COVID Symptom Study Biobank Consortium, J. D. Carpentieri, Claire J. Steves

**Author notes:** A list of authors and their affiliations are given below. **The COVID Symptom Study Biobank Consortium**. Michela Antonelli^1^, Vicky Bowyer^2^, Julia Brown^2,3^, Liane Canas^1^, Joan Capdevila Pujol^4^, Nathan Cheetham^2^, Lynn Cherkas^2^, Jie Deng^1^, Katie Doores^5^, Emma Duncan^2,6^, Maria Paz Garcia^2^, Alexander Hammers^1,7^, Deborah Hart^2^, Nicholas Harvey^2^, Adrian Hopper^8^, Christina Hu^4^, Eric Kerfoot^1^, Michael Malim^5^, Marc Modat^1^, Erika Molteni^1^, Benjamin Murray^1^, Ayrun Nessa^2^, Sebastien Ourselin^1^, Tim Spector^2^, Claire Steves^2,9^, Carole Sudre^1,10,11^, Samuel Wadge^2^, Jonathan Wolf^4^. School of Biomedical Engineering & Imaging Sciences, King’s College London, London, United Kingdom. Department of Twin Research and Genetic Epidemiology, King’s College London, London, United Kingdom. Zoe Ltd, 164 Westminster Bridge Road, London, United Kingdom. Department of Infectious Diseases, King’s College London, London, United Kingdom. Department of Endocrinology, Guy’s and St Thomas’ NHS Foundation trust, London, United Kingdom. Guy’s and St Thomas’ NHS Foundation trust, London, United Kingdom. Department of Ageing and Health, Guy’s and St Thomas’ NHS Foundation trust, London, United Kingdom. MRC Unit for Lifelong Health and Ageing, Department of Population Science and Experimental Medicine, University College London, London, United Kingdom. Centre for Medical Image Computing, Department of Computer Science, University College London, London, United Kingdom.

## Abstract

**Introduction:** Inequalities in health care access and experiences during the COVID-19 pandemic have been observed along axes of social advantage. It is less well understood how access to care varied intersectionally with combinations of multiple social factors, and how social advantage shaped care experiences for COVID-19 illness.

**Methods:** We analysed responses to both closed (N = 3,516) and open (N = 335) survey questions relating to health and social care access and experiences during the first two and a half years of the COVID-19 pandemic in the United Kingdom community-based cohort, COVID Symptom Study Biobank. Causal effects of individual socio-demographic variables on access to health and social care were estimated with multivariable regression models, weighted for inverse probability of survey completion and adjusted for potential confounders. Associations between care access issues and social strata comprising combinations of sex, education level and local area deprivation were estimated using the intersectional multilevel analysis of individual heterogeneity and discriminatory accuracy (MAIHDA) approach. Responses to open questions on health care experiences for COVID-19 illness were deductively coded and quantitatively analysed to estimate associations between socio-demographic advantage and various aspects of care experiences.

**Results:** Gradients in health and social care access along the lines of social advantage were observed in intersectional MAIHDA models, with the predicted probability of access issues highest for the stratum comprising female participants with lowest education and highest deprivation levels (42.9%, 95% CI: 31.9%-54.6%), and lowest for male participants with highest education and lowest deprivation (18.7%, 95% CI: 12.8%-26.7%). Socially disadvantaged participants also reported receiving poorer care for COVID-19, with lower likelihood of reporting receiving adequate care and specialist care for long COVID, and higher likelihood of negative experiences of care vs. advantaged participants.

**Conclusions:** Inequalities in likelihood of health and social care access issues were observed, as well as inequalities in care experiences specifically for COVID-19, with issues accessing care and poorer experiences more likely to be reported by individuals with greater social disadvantage.

**Funding:** Chronic Disease Research Foundation, National Institute for Health and Care Research, Medical Research Council, Zoe Ltd.

**Key terms:** Patient experience; Health care access; Quality of care; Long COVID; Mixed methods; Intersectionality

**Patient or Public Contribution:** Members of the COVID Symptom Study Biobank Volunteer Advisory Panel provided feedback on the questionnaire that was our primary data source for this analysis. Previous discussions with the Volunteer Advisory Panel informed the aims and objectives of this study.

**Plain language summary:** Problems getting care for health issues has been common in the COVID-19 pandemic. Problems have also been more common for certain groups, such as women, people from ethnic minorities, and people in lower paid jobs. But there hasn’t been much research that has looked at how problems getting care has varied between people with different combinations of characteristics. Research also hasn’t looked very much at whether people with different circumstances had different experiences of getting care for COVID-19. In this study, we looked at responses to a questionnaire from August 2022 from a health study focused on understanding the long term effects of COVID-19. We found that our female participants tended to be more likely to experience problems accessing health and social care services. We also found that participants with disadvantaged circumstances, such as having a low education level, living in a deprived area, and having low income, were more likely to have problems accessing care. Similarly, participants with more disadvantaged circumstances had negative experiences of getting health care for COVID-19 more often than more advantaged participants. These results suggest that health care access and experiences vary significantly depending on people’s circumstances. This differences in experiences need to be addressed, particularly for people living with symptoms of long COVID.

## Introduction

Disruptions to health and social care services during the COVID-19 pandemic have been commonly reported in the United Kingdom [1,2], and have been associated with lower probability of self-reported recovery from COVID-19 [3]. Issues accessing care during the pandemic appear to have been unequally distributed along axes of structural social advantage, with higher likelihood of disruptions for female vs. male sex, racially minoritised vs. white ethnic groups, and for those in manual/routine vs. managerial/professional occupations. Gradients in health and health outcomes have also been consistently observed over time within the UK, including in the most recent Census in 2021, along similar axes of advantage including race/ethnicity [4], education level [5,6], occupational class [7,8], income [9], and local area deprivation [10]. Social gradients were also seen for COVID-19 incidence and mortality [11,12], and prevalence and recovery from long-term ongoing symptoms, or long COVID [3,13,14], with poorer outcomes for those with greater social disadvantage.

Discrepancies between prevalence of long COVID based on self-report [15–17], and from coding in electronic health care records (EHR) in the UK [18–20], where prevalence is highest for those in the most deprived areas by self-report, but coded in EHR most often for those in the least deprived areas, as well as greater symptom intensity among more disadvantaged groups within long COVID clinical populations [21], suggests a greater unmet need for long COVID care within more disadvantaged populations. In combination, these reports suggest the presence of the ‘inverse care law’ [22], during the COVID-19 pandemic, where the availability of health care was associated inversely with health care need.

In qualitative studies of health care access and experiences of those living with long COVID in England, Scotland, and Austria, common themes include difficulties getting appointments with general practitioners, struggling to navigate complex health care systems, lack of awareness and knowledge of long COVID from health care providers, anticipated stigma based on minoritised identity factors or stereotyping of long COVID, and negative, often discriminatory, experiences with health care providers [23–29]. Intersectional inequalities have also been observed during the COVID-19 pandemic in both access to care [30], and in experiences of care [23,29], with greater access issues and poorer experiences for women from racially minoritised or immigrant backgrounds. However, no studies have looked at socio-demographic variation in health care access and experiences using a quantitative intersectional approach. To-date, outcomes in quantitative studies have been limited to access to health care, and examined socio-demographic factors as individual exposures only. There have also been no studies that have considered responses to open and closed survey questions in combination.

Given reports of health care access and experiences being shaped by explicit combinations or ‘intersections’ of socio-demographic factors in smaller qualitative studies, there is a desire for more research to understand whether quantitative, intersectional differences in care access and experiences are replicated within larger samples [31]. Furthermore, understanding socio-demographic variation in care specifically for COVID-19 is also particularly crucial, given the known impacts of COVID-19 and long COVID on health and wider circumstances [32], and the effectiveness of early pharmacological treatment in reducing the likelihood of long COVID [33], and of specialist care programs taking a cognitive and behavioural approach for those living with long COVID [34].

Therefore, in this study, we use a mixed methods approach to answer the research question: how did pre-pandemic social advantage and disadvantage effect access to, and experiences of, health and social care during the COVID-19 pandemic? Firstly, we estimated associations between pre-pandemic markers of social advantage and disadvantage and health and social care access issues/disruptions during the first two years of the COVID-19 pandemic using self-reported responses to closed survey questions. We estimated associations with socio-demographic factors both individually and in combination using an intersectional approach. Secondly, we qualitatively coded a subset of responses to an open-text question where participants described experiences of health care specifically for COVID-19 illness; we then quantitatively analysed how care access and experiences varied according to cumulative social advantage or disadvantage, while accounting for health care need. Quantitative analyses of coded open-text responses are supported by illustrative quotations. We hypothesised that social strata with combinations of advantaged positions/statuses would be less likely to report experiencing access issues and report better experiences of care, while those with combinations of multiple disadvantaged statuses would be more likely to report access issues and worse care experiences.

## Methods

### Data sources

#### COVID Symptom Study Biobank

Study participants were volunteers from the COVID Symptom Study Biobank (CSSB) cohort. CSSB participants were recruited via the COVID Symptom Study app from ZOE Ltd (CSS) launched in the UK on March 24, 2020. All data were collected with informed consent obtained online. The CSS app allowed participants to self-report demographic information, symptoms potentially suggestive of COVID-19 infection, any SARS-CoV-2 testing and results, and any vaccinations. CSS participants from across the UK were invited to join the CSSB by email in October to November 2020 and May to June 2021. Invitation to join the CSSB targeted five groups with different statuses at the time of invitation, derived from CSS data: asymptomatic COVID (positive SARS-CoV-2 test and no associated symptoms); “short COVID” (positive SARS-CoV-2 test and 1-13 days of symptoms); “long COVID” (positive SARS-CoV-2 test and ≥ 28 days’ symptoms); “long non-COVID” (negative SARS-CoV-2 test and ≥ 28 days’ symptoms); and “healthy non-COVID” (negative SARS-CoV-2 test and ≤ 3 days with ≤ 3 symptoms). Before invitation, individuals were matched based on minimum Euclidian distance for age, sex and body mass index (BMI) across groups. The aim of the targeted approach was to give five equally-sized, matched groups, to allow investigation of the long-term effects of COVID-19 on health. As such, cohort composition in terms of prevalence of COVID-19 and long COVID at the time of recruitment is not representative of the wider UK population.

CSSB participants were invited (N = 8324) to participate in the *“COVID Reflections - Two Years On”* online questionnaire in August 2022. Questionnaire data was supplemented with data collected at time of registration with the CSS app, at consent to CSSB, and in earlier CSSB studies. Variables described below were collected as part of the August 2022 questionnaire unless otherwise stated. The CSSB Volunteer Advisory Panel were consulted on the delivery of the August 2022 questionnaire and gave recommendations on invitation and reminder strategies.

#### Outcome variables (closed question analysis): Health and social care access issues during the COVID-19 pandemic

Health and social care access issues during the COVID-19 pandemic, independent of reason for seeking care, were measured in the August 2022 CSSB questionnaire as part of a more general question on adverse pandemic experiences asked to all questionnaire respondents, *“Have you experienced any of the following during the COVID-19 pandemic?”*. Respondents were asked to report Yes/No/Prefer not to answer for the following items: *a. Lost your job/been unable to do paid work, b. Been put on furlough from your job (paid leave), c. Unable to pay bills, d. Evicted/lost accommodation, e. Unable to afford food, f. Unable to access required medication, g. Unable to access health services in the community, for instance, GP/community physiotherapy/nurse/podiatrist/dentist, h. Unable to access the community social care services or voluntary sector support you need, for instance, from carers or day centres, i. Unable to access inpatient or outpatient appointments booked at a hospital for a consultation, investigation, treatment, or surgery, j. Unable to access appointment for cognitive behaviour therapy, counselling, or psychological therapy, k. You lost somebody close to you due to COVID-19, l. Change in relationship status*. Five response items, *f*, *g*, *h*, *i*, and *j*, were used to derive a continuous variable counting the number of health and social care access issues, and a binary variable indicating the presence of one or more issues.

#### Outcome variables (open question analysis): Health care experiences for COVID-19 illness

Outcome variables describing aspects of health care access and quality, specifically for COVID-19 illness, were derived from qualitative coding, described in detail further on, of responses to the open-text question, *“Please use the space below to tell us a bit more about any care you received for COVID-19? For example, was it easy to see your GP? Were there any parts of your treatment that were particularly helpful or that you think could be improved? Please write as much or as little as you like about any aspects of your healthcare experience.”*, prefaced with, *“We are interested in hearing about your experience with the healthcare system and how those experiences have impacted your health and, if relevant, any COVID-19 symptoms.”*.

To generate variables for quantitative analysis, qualitative coding of open-text responses was done by two of the authors using a deductive approach, using a predetermined codebook generated based on the research questions of the study (full codebook in with unadjusted frequencies in Table S5) [35]. Outcome variables were generated from either single codes or combinations of multiple codes (Table 1).

**Table 1.** Qualitative codes used to define COVID-19 health care outcomes for quantitative analysis. Full codebook is given in Table S5.

| Outcome: Category | Qualitative code(s) description | Code # |
| --- | --- | --- |
| Care needed for COVID-19: Yes | Did not receive care for COVID-19 -> did not attempt to contact health care services -> but wanted or needed to | 11 |
|  | OR<br>Did not receive care for COVID-19 -> Attempted to use health care services but did not receive care | 13 |
|  | OR<br>Received care for COVID-19 | 16 |
| Care needed for COVID-19: No | Did not receive care for COVID-19 -> did not attempt to contact health care services -> and did not need them | 12 |
| Care needed for COVID-19: Unknown | None of the above “Care needed for COVID-19” codes |  |
| Care received: Yes, inferred as adequate | Received care for COVID-19<br>AND NOT coded with any of the below “Care received” codes | 16 |
| Care received: Yes, perceived as inadequate | Received care for COVID-19 -> Received care (perceived as) inadequate - care received was unhelpful or irrelevant, had appointment but effectively no care provided, etc. | 25 |
| Care received: No, did not attempt to access | Did not receive care for COVID-19 -> did not attempt to contact health care services -> but wanted or needed to | 11 |
| Care received: No, access attempt unsuccessful | Did not receive care for COVID-19 -> Attempted to use health care services but did not receive care | 13 |
| Received care from long COVID clinic or specialist | Received care for COVID-19 -> Received care in 'long covid clinic' or from specialist | 19 |
| Ease of care access: Easy | Received care for COVID-19 -> Accessing was easy/straightforward (explicit or inferred) | 21 |
| Ease of care access: Difficult | Received care for COVID-19 -> Accessing was difficult (explicit or inferred) | 22 |
| Ease of care access: Did not attempt access | Did not receive care for COVID-19 -> did not attempt to contact health care services -> but wanted or needed to | 11 |
| Ease of care access: Not stated | None of the above "Ease of care access" codes |  |
| Perception of care experience: Positive | Positive experience | 1 |
| Perception of care experience: Negative | Negative experience | 6 |
| Perception of care experience: Neutral or mixed | Neutral/mixed | 4 |
| Perception of care experience: Not clear | Unclear/not possible to discern<br>OR<br>no code assigned | 5 |

The approach was not completely prescriptive, with several additional codes developed during the coding process to ensure the codebook captured the breadth of the data [36]. To ensure consistency of coding, coders first met to develop a shared theoretical understanding, interpretation, and application of each code within the codebook before starting coding. To allow a closer reading of the data in relation to each topic, the codebook was split into three thematic framings: ‘Reflection on health care experiences’; ‘Description of healthcare experiences’; and ‘Mention of influences that related to or affected experiences with care’. A round of coding was undertaken for each theme. After independently coding each thematic section, coders came together to compare and compile codes.

Any divergence in coding was resolved through discussion and often tended towards a consensus around one coders’ interpretation. If consensus was not reached via this discursive approach, the open-text response in question would be analysed by both coders and a compromise reached that both were comfortable with [37]. For assessment of whether care received was adequate or inadequate, an absence of explicit mention of dissatisfaction or the need for further care in open-text responses was ‘*inferred* as adequate’, while those where care was ‘*perceived* as inadequate’ were based on explicit mentions in the response. Assessment of ease of care access was based on inference from the coders based on the text, as well as explicit mentions by the respondent (see Table 5 quote 11 as an example where easy access was inferred, quote 5 as example of explicit mention of difficult access).

#### Socio-demographic, health and COVID-19 illness characteristics

Socio-demographic characteristics were derived from self-report at registration to the CSS app, CSSB consent, or in the August 2022 CSSB questionnaire. Health characteristics were derived from self-report at registration to the CSS app, CSSB consent, from a February 2021 CSS questionnaire, or from May 2021 and August 2022 CSSB questionnaires. COVID-19 illness characteristics were derived from self-report in the August 2022 questionnaire. Further details relating to data sources, question wording and processing prior to analysis are given in Supplementary Information Section S1.

### Eligibility criteria and sample selection

For all analyses, inclusion criteria were complete data on age, sex, ethnic group, and area of residence.

For analysis of health and social care access issues during the COVID-19 pandemic from closed question responses, individuals with partial completion of the 2022 CSSB questionnaire were excluded, giving an analysis sample of N = 3,516. A full sample selection flow diagram is given in Figure S1.

For analysis of health care experiences for COVID-19 from open text responses, sample selection was designed to allow the effect of multiple pre-pandemic advantage/disadvantage to be estimated. To limit the number of open text responses to qualitatively code by hand to a manageable level, a subsample of all respondents with symptomatic SARS-CoV-2 infection and whose longest symptom duration COVID-19 episode started at least 84 days before questionnaire completion was chosen. Prior to subsampling, participants were categorised into three socio-demographic groups representing the accumulation of advantaged or disadvantaged statuses in the context of systems of social power and axes of inequity [38], named ‘relatively advantaged’, ‘intermediate’, and ‘relatively disadvantaged’. To generate socio-demographic groups, categories of ethnic group, education level, local area deprivation, employment status and household income were first ascribed as representing relatively advantaged or disadvantaged statuses, as shown in Table 2. Socio-demographic groups were then derived by categorisation of the number of advantages minus the number of disadvantages, as shown in Table 3. Participants with values of advantages-disadvantages = 0 and +2 were not included to create clear demarcations between groups. Finally, subsets of each socio-demographic group were selected using stratified random sampling, with subsets proportionally representative in terms of age group, sex, long COVID status and self-perceived COVID-19 recovery status. The fraction of the socio-demographic group selected for each subset was varied to give three subsets of approximately equal size. Aside from household income, which was collected in the 2022 questionnaire, other factors represent pre-pandemic status. While gender was hypothesised as an additional axis of inequity, sex (as a proxy for gender) was not used to generate socio-demographic groups, as it was thought more important that each group contained responses from female and male participants.

**Table 2.** Categories ascribed as representing relative advantage or disadvantage within systems of social power and axes of inequity, in the derivation of socio-demographic groups representing accumulation of advantaged or disadvantaged statuses.

| **Factor** | **Advantaged status** | **Disadvantaged status** |
| --- | --- | --- |
| Ethnic group | White groups | Racially minoritised groups (Asian/Asian British, Black/Black British, Mixed/multiple, Any other) |
| Education level | Postgraduate level | Less than degree level |
| Local area deprivation | Low deprivation area (Index of Multiple Deprivation (IMD) Decile 8-10) | High deprivation area (IMD Decile 1-3) |
| Pre-pandemic employment status | - | Unemployed or long-term sick or disabled |
| Annual gross household income | High (£100,000 or higher) | - |

**Table 3.** Sample sizes and values for the number of advantages minus the number of disadvantages of socio-demographic groups used in analysis of open text question on COVID-19 health care experiences.

| **Socio-demographic group** | **#Advantages-#Disadvantages** | **Sample size of qualitatively coded subset** |
| --- | --- | --- |
| Relatively advantaged | +3, +4 | 109 |
| Intermediate | +1 | 117 |
| Relatively disadvantaged | -1, -2, -3 | 109 |

### Statistical analysis

#### Regression models & proposed causal pathways

To obtain estimates of the effects of individual socio-demographic variables on the number and likelihood of reporting health and social care access issues during the COVID-19 pandemic, we used multivariable ordinary least squares (OLS) linear regression models (continuous outcome variable) and poisson regression models with robust errors (binary outcome variable) [39]. Models used the “HC3” estimator of coefficient standard errors to account for heteroskedasticity [40].

For each exposure variable of interest, separate models were run including potential confounding variables as appropriate, based on the hypothesised directed acyclic graph (DAG), developed using DAGitty software: http://www.dagitty.net/dags.html (abridged DAG in Figure 1, full DAG in Figure S2, full DAGitty code given in supplementary information)[41].

**Figure 1.**
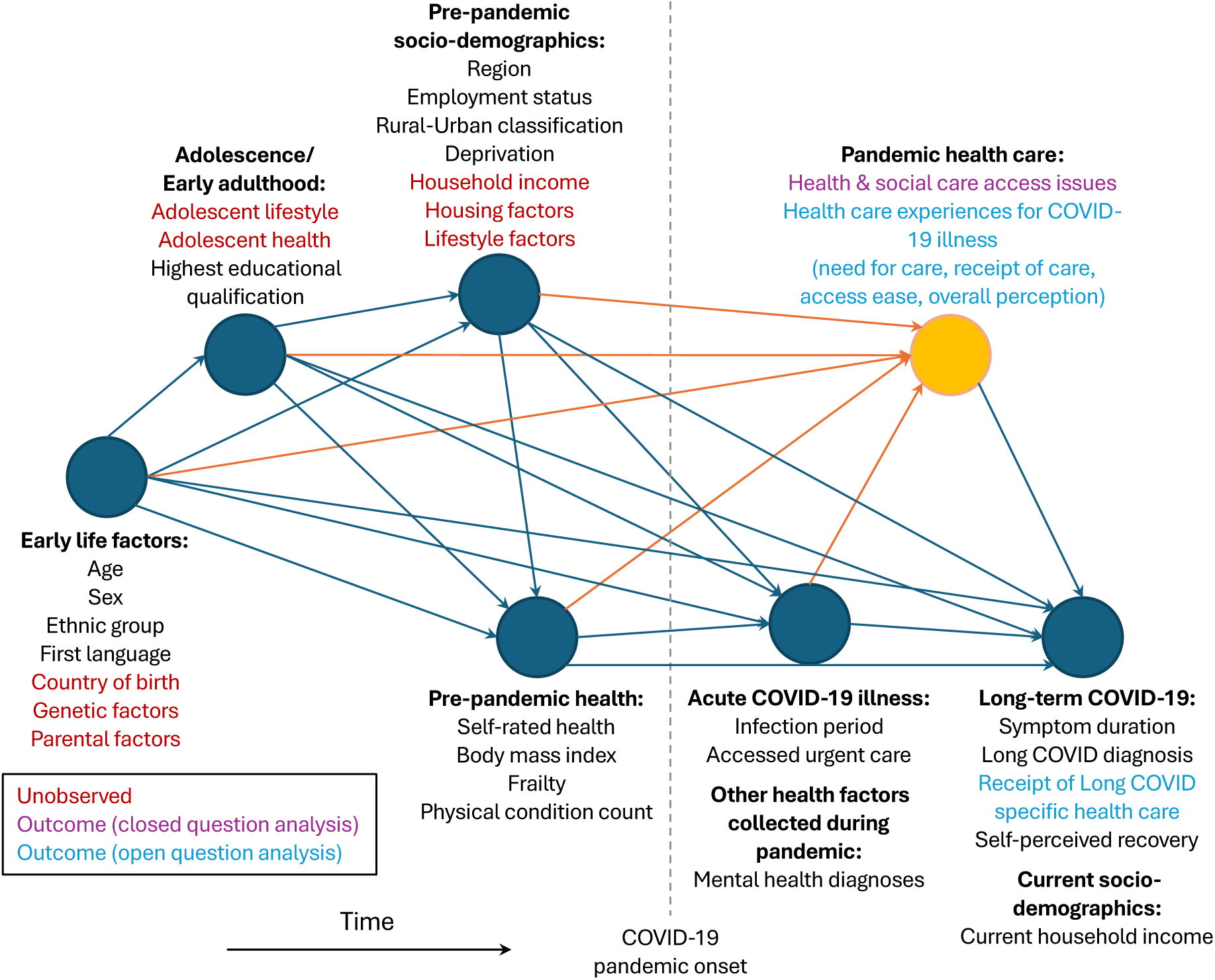
Directed acyclic graph describing hypothesised causal pathways. Proposed directed acyclic graph (DAG) used to generate minimal adjustment variable sets for models estimating the association between exposure variables and outcomes related to health care during the COVID-19 pandemic. Outcome variables derived from closed and open survey questions are highlighted in purple and blue respectively, while key unobserved potential confounders are coloured in red. The DAG is structured in order of hypothesised data generation/crystallisation from left to right, and variables with similar time of data generation/crystallisation are grouped for clarity into ‘super nodes’. The pandemic health care super node and direct arcs to it are coloured in orange, to distinguish the node containing the majority of outcomes examined. The proposed DAG is ‘saturated’, in that each variable is hypothesised to be caused by all earlier variables.

Associations between experiencing health and social care access issues and combinations of sex, education level and local area deprivation were estimated using mixed-effects linear and logistic regression models, following the intersectional multilevel analysis of individual heterogeneity and discriminatory accuracy (MAIHDA) approach [42,43], informed by the intersectionality framework [44,45]. In models, a ‘social strata’ variable detailing the explicit combination of sex, education and deprivation acted as a level 2, random (intercept) effects variable, while the individual variables of sex, education and deprivation, plus age group and ethnic group as potential confounders, were included as level 1 fixed effects. Sex was used as a proxy for gender identity based on data availability. Sex, education, and deprivation were chosen because of associations with sex and socio-economic status previously observed in analyses of individual factors [1,2], while also having strata sizes large enough to make reliable estimates.

For each social stratum, we present the stratum-level average predicted number of access issues (from linear model) or the average predicted probability of one or more issues (from logistic model) generated from the fitted models, which capture both additive and interaction effects. The R script used to fit MAIHDA models was adapted from a MAIHDA tutorial [42,46]. 95% confidence intervals for predicted probabilities are approximate only because the model assumes no sampling covariability between the regression coefficients and stratum random effects.

As supplementary and sensitivity analyses, additional MAIHDA models were run where: 1) the analysis sample was stratified by COVID-19 history, to test how access to care varied within subsets of respondents with similar experiences of COVID-19 illness; 2) household income, at the time of the 2022 questionnaire, was also included in the social strata variable, to further test the role of economic capital; 3) pre-pandemic health factors were included as additional potential confounders, to test an alternative data generating timeline where health factors predominantly precede socio-demographic factors.

To estimate the association between socio-demographic group and aspects of COVID-19 health care experiences, in analysis of qualitatively coded open text responses, multivariable poisson regression models were again used, with age group included as an adjustment variable.

#### Generation of inverse probability weights

Regression models included weights representing participants’ inverse probability of questionnaire response and selection into the given analysis sample. Weights were used to reduce potential selection bias from differential response rates along socio-demographic lines, as is often seen with volunteer cohorts [47], as well as collider bias from conditioning on COVID-19 infection for analysis of COVID-19 health care experiences. If infection is a mediating factor between exposures (pre-pandemic socio-demographics) and outcome (health care experiences), any unmeasured factor associated with both COVID-19 infection and our outcome may bias estimated associations [48,49]. Sets of weights were generated following a forward sequential feature selection method that optimised prediction (area under the receiver operating characteristic curve, AUC-ROC, score) of presence in the analysis sample, described in more detail in previous CSSB studies [3,50]. Weights were winsorised (bottom and top 5% values set equal to 5^th^ and 95^th^ percentile values), to limit influence of individuals with extreme weight values, prior to use in regression models. Details of models used to generate inverse probability weights (IPWs) are summarised in Table S1.

#### Software

Fixed-effects regression models were fit, and figures were created, using python v3.8.8 and packages: numpy v1.20.1, pandas v1.2.4, statsmodels v0.12.2, scipy v1.6.2, scikit-learn v0.24.1, matplotlib v3.3.4, seaborn v0.11.1. Intersectional MAIHDA mixed-effects regression models were fit using R v4.3.0 and packages: haven v2.5.4, tidyverse v2.0.0, ggeffects v1.5.2, lme4 v1.1.35.2, merTools v0.6.2, labelled v2.13.0, sjPlot v2.8.16, Metrics v0.1.4, dplyr v1.1.4.

## Results

### Part 1: Health and social care access issues during the COVID-19 pandemic (closed question analysis)

#### Sample characteristics

Analysis of health and social care access issues during the COVID-19 pandemic from closed questions was based on 3,516 participants who completed the 2022 CSSB questionnaire (of 8,324 invited, 42% response rate, sample selection shown in Figure S1). The median age group of respondents was 50-59 years old, most were female sex (81%), and identified as of white ethnic groups (97%) (Table 4). Respondents were relatively socio-economically advantaged in comparison to the general UK population, tending to have high education and income levels, living in lower deprivation areas, and were mostly employed at the beginning of the COVID-19 pandemic. Issues accessing health and social care were common, with 28% reporting one or more of the five issue types, and an average of 0.44 issues reported across all participants. The most common issue was accessing community based services (24%), and least common was access to social care (2%). Among those reporting one or more care access issues, most experienced a single issue only (62%), with smaller proportions reporting two or more (38%), and three or more (12%) issues.

**Table 4.** Socio-demographic characteristics and health and social care access issues. Unweighted group sizes, unadjusted proportions of participants with one or more health and social care access issues during the COVID-19 pandemic, and mean number of issues, split by participant socio-demographics. Socio-demographic categories acting as references in multivariable regression models are indicated. Figures for group sizes of less than 5 are suppressed.

| Domain | Variable | Group size |  | One or more health & social care access issues |  | Mean number of issues |
| --- | --- | --- | --- | --- | --- | --- |
|  |  | N | % | N | % |  |
|  | **Total** | 3516 |  | 999 | 28.4% | 0.44 |
| **Health & social care access issues during the COVID-19 pandemic** | **Health & social care access issue type** |  |  |  |  |  |
|  | Unable to access required medication | 144 | 4.1% |  |  |  |
|  | Unable to access health services in the community | 827 | 23.5% |  |  |  |
|  | Unable to access the community social care services or voluntary sector support needed | 73 | 2.1% |  |  |  |
|  | Unable to access inpatient or outpatient appointments booked at a hospital | 388 | 11.0% |  |  |  |
|  | Unable to access appointment for cognitive behaviour therapy, counselling, or psychological therapy | 127 | 3.6% |  |  |  |
|  | **Health & social care access issue count** |  |  |  |  |  |
|  | None | 2517 | 71.6% |  |  |  |
|  | One | 624 | 17.7% |  |  |  |
|  | Two | 251 | 7.1% |  |  |  |
|  | Three | 81 | 2.3% |  |  |  |
|  | Four | 25 | 0.7% |  |  |  |
|  | Five | 18 | 0.5% |  |  |  |
| **Individual pre-pandemic demographics** | **Age (median, interquartile range)** | 59 (52-65) |  | 58 (52-65) |  |  |
|  | **Age group (years)** |  |  |  |  |  |
|  | 18-39 | 186 | 5.3% | 47 | 25.3% | 0.51 |
|  | 40-49 | 493 | 14.0% | 148 | 30.0% | 0.47 |
|  | 50-59 (reference) | 1144 | 32.5% | 362 | 31.6% | 0.51 |
|  | 60-69 | 1254 | 35.7% | 322 | 25.7% | 0.37 |
|  | ≥ 70 | 439 | 12.5% | 120 | 27.3% | 0.40 |
|  | **Sex** |  |  |  |  |  |
|  | Female (reference) | 2840 | 80.8% | 843 | 29.7% | 0.47 |
|  | Male | 676 | 19.2% | 156 | 23.1% | 0.32 |
|  | **Ethnic group** |  |  |  |  |  |
|  | Asian/Asian British | 18 | 0.5% | <5 | - | 0.33 |
|  | Black/Black British | 15 | 0.4% | <5 | - | 0.27 |
|  | Mixed/Multiple | 29 | 0.8% | 9 | 31.0% | 0.45 |
|  | Other | 38 | 1.1% | 11 | 28.9% | 0.42 |
|  | White (reference) | 3416 | 97.2% | 972 | 28.5% | 0.44 |
|  | **First language** |  |  |  |  |  |
|  | English (reference) | 3437 | 97.8% | 976 | 28.4% | 0.44 |
|  | Other | 76 | 2.2% | 23 | 30.3% | 0.50 |
|  | Prefer not to answer/not stated | 3 | 0.1% | 0 | 0.0% | 0.00 |
|  | **Highest educational qualification** |  |  |  |  |  |
|  | Prefer not to answer/not stated | 40 | 1.1% | 13 | 32.5% | 0.58 |
|  | Did not complete secondary school | 25 | 0.7% | <5 | - | 0.28 |
|  | GCSE or GNVQ or equivalent | 412 | 11.7% | 139 | 33.7% | 0.52 |
|  | A-Levels or advanced GNVQ or equivalent | 587 | 16.7% | 183 | 31.2% | 0.45 |
|  | University degree (reference) | 1269 | 36.1% | 341 | 26.9% | 0.44 |
|  | Postgraduate degree or higher | 1014 | 28.8% | 272 | 26.8% | 0.42 |
|  | PhD | 169 | 4.8% | 47 | 27.8% | 0.43 |
|  | **UK Region** |  |  |  |  |  |
|  | East Midlands | 227 | 6.5% | 74 | 32.6% | 0.56 |
|  | East of England | 377 | 10.7% | 113 | 30.0% | 0.49 |
|  | London (reference) | 593 | 16.9% | 163 | 27.5% | 0.45 |
|  | North East | 109 | 3.1% | 29 | 26.6% | 0.41 |
|  | North West | 342 | 9.7% | 95 | 27.8% | 0.40 |
|  | Scotland & Northern Ireland | 178 | 5.1% | 54 | 30.3% | 0.50 |
|  | South East | 693 | 19.7% | 195 | 28.1% | 0.44 |
|  | South West | 360 | 10.2% | 90 | 25.0% | 0.41 |
|  | Wales | 180 | 5.1% | 53 | 29.4% | 0.44 |
|  | West Midlands | 223 | 6.3% | 67 | 30.0% | 0.44 |
|  | Yorkshire and The Humber | 234 | 6.7% | 66 | 28.2% | 0.37 |
|  | **Pre-pandemic employment status** |  |  |  |  |  |
|  | Employed (reference) | 1777 | 50.5% | 530 | 29.8% | 0.45 |
|  | Self-employed | 390 | 11.1% | 106 | 27.2% | 0.42 |
|  | Unemployed | 15 | 0.4% | 6 | 40.0% | 0.87 |
|  | Permanently or long-term sick or disabled | 46 | 1.3% | 35 | 76.1% | 1.83 |
|  | Retired | 928 | 26.4% | 206 | 22.2% | 0.32 |
|  | Other | 287 | 8.2% | 87 | 30.3% | 0.48 |
|  | Unknown | 73 | 2.1% | 29 | 39.7% | 0.78 |
|  | **Rural-Urban classification** |  |  |  |  |  |
|  | Rural | 864 | 24.6% | 227 | 26.3% | 0.39 |
|  | Urban (reference) | 2652 | 75.4% | 772 | 29.1% | 0.46 |
|  | **Local area deprivation** |  |  |  |  |  |
|  | IMD Decile 1 (most deprived 10%) | 82 | 2.3% | 37 | 45.1% | 0.73 |
|  | IMD Decile 2 | 129 | 3.7% | 42 | 32.6% | 0.52 |
|  | IMD Decile 3 | 185 | 5.3% | 57 | 30.8% | 0.50 |
|  | IMD Decile 4 | 255 | 7.3% | 90 | 35.3% | 0.58 |
|  | IMD Decile 5 | 314 | 8.9% | 95 | 30.3% | 0.49 |
|  | IMD Decile 6 | 406 | 11.5% | 122 | 30.0% | 0.48 |
|  | IMD Decile 7 | 428 | 12.2% | 129 | 30.1% | 0.48 |
|  | IMD Decile 8 | 495 | 14.1% | 131 | 26.5% | 0.41 |
|  | IMD Decile 9 | 563 | 16.0% | 150 | 26.6% | 0.41 |
|  | IMD Decile 10 (least deprived 10%) (reference) | 659 | 18.7% | 146 | 22.2% | 0.31 |
| **Socio-demographics during pandemic** | **Current household income** |  |  |  |  |  |
|  | Prefer not to answer/not stated | 467 | 13.3% | 146 | 31.3% | 0.49 |
|  | Less than £20,000 | 318 | 9.0% | 127 | 39.9% | 0.68 |
|  | £20,000-£29,999 | 348 | 9.9% | 103 | 29.6% | 0.47 |
|  | £30,000-£39,999 | 425 | 12.1% | 128 | 30.1% | 0.48 |
|  | £40,000-£49,999 | 415 | 11.8% | 111 | 26.7% | 0.46 |
|  | £50,000-£74,999 (reference) | 649 | 18.5% | 178 | 27.4% | 0.40 |
|  | £75,000-£99,999 | 378 | 10.8% | 106 | 28.0% | 0.41 |
|  | £100,000 or more | 516 | 14.7% | 100 | 19.4% | 0.28 |

**Table 5.**
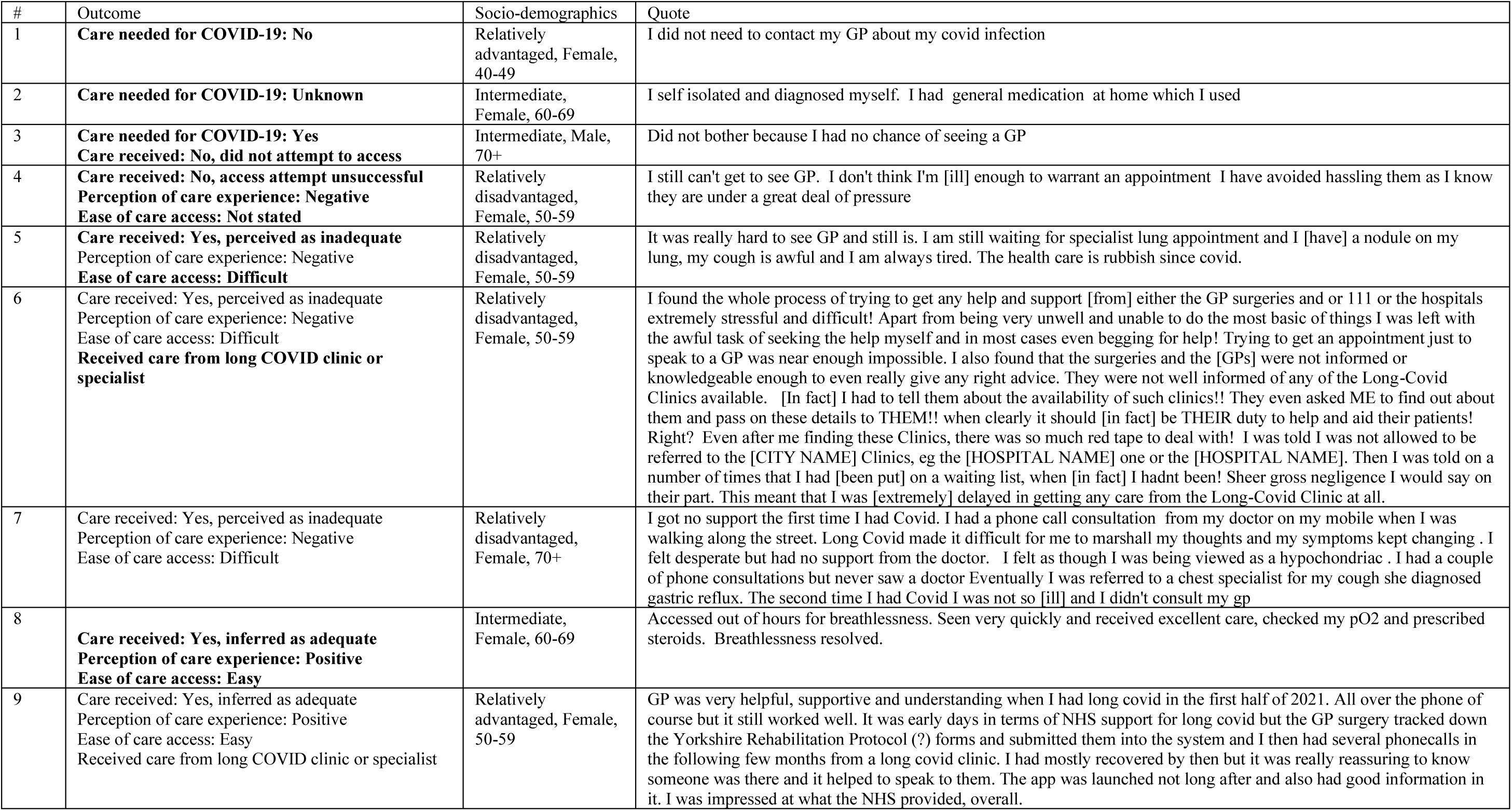
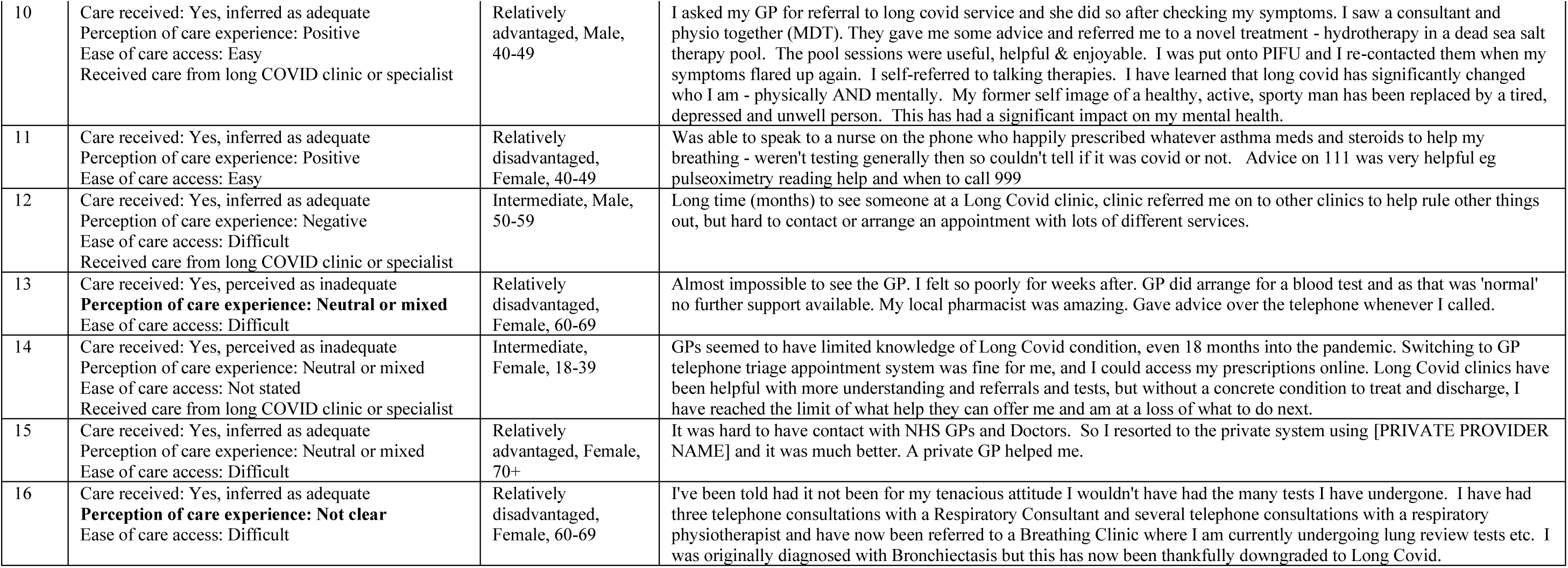
Illustrative quotes from free-text responses describing COVID-19 health care experiences, including the health care outcomes ascribed to the quotes, socio-demographic group used in quantitative analysis, sex and age group. The first quote that includes a certain type of experience is highlighted in bold. Potentially identifying text is masked and spelling is occasionally edited for clarity. Quotes are otherwise reproduced verbatim.

| # | Outcome | Socio-demographics | Quote |
| --- | --- | --- | --- |
| 1 | **Care needed for COVID-19: No** | Relatively advantaged, Female, 40-49 | I did not need to contact my GP about my covid infection |
| 2 | **Care needed for COVID-19: Unknown** | Intermediate, Female, 60-69 | I self isolated and diagnosed myself. I had general medication at home which I used |
| 3 | **Care needed for COVID-19: Yes**<br>**Care received: No, did not attempt to access** | Intermediate, Male, 70+ | Did not bother because I had no chance of seeing a GP |
| 4 | **Care received: No, access attempt unsuccessful**<br>**Perception of care experience: Negative**<br>**Ease of care access: Not stated** | Relatively disadvantaged, Female, 50-59 | I still can't get to see GP. I don't think I'm [ill] enough to warrant an appointment I have avoided hassling them as I know they are under a great deal of pressure |
| 5 | **Care received: Yes, perceived as inadequate**<br>Perception of care experience: Negative<br>**Ease of care access: Difficult** | Relatively disadvantaged, Female, 50-59 | It was really hard to see GP and still is. I am still waiting for specialist lung appointment and I [have] a nodule on my lung, my cough is awful and I am always tired. The health care is rubbish since covid. |
| 6 | Care received: Yes, perceived as inadequate<br>Perception of care experience: Negative<br>Ease of care access: Difficult<br>**Received care from long COVID clinic or specialist** | Relatively disadvantaged, Female, 50-59 | I found the whole process of trying to get any help and support [from] either the GP surgeries and or 111 or the hospitals extremely stressful and difficult! Apart from being very unwell and unable to do the most basic of things I was left with the awful task of seeking the help myself and in most cases even begging for help! Trying to get an appointment just to speak to a GP was near enough impossible. I also found that the surgeries and the [GPs] were not informed or knowledgeable enough to even really give any right advice. They were not well informed of any of the Long-Covid Clinics available. [In fact] I had to tell them about the availability of such clinics!! They even asked ME to find out about them and pass on these details to THEM!! when clearly it should [in fact] be THEIR duty to help and aid their patients! Right? Even after me finding these Clinics, there was so much red tape to deal with! I was told I was not allowed to be referred to the [CITY NAME] Clinics, eg the [HOSPITAL NAME] one or the [HOSPITAL NAME]. Then I was told on a number of times that I had [been put] on a waiting list, when [in fact] I hadn't been! Sheer gross negligence I would say on their part. This meant that I was [extremely] delayed in getting any care from the Long-Covid Clinic at all. |
| 7 | Care received: Yes, perceived as inadequate<br>Perception of care experience: Negative<br>Ease of care access: Difficult | Relatively disadvantaged, Female, 70+ | I got no support the first time I had Covid. I had a phone call consultation from my doctor on my mobile when I was walking along the street. Long Covid made it difficult for me to marshall my thoughts and my symptoms kept changing. I felt desperate but had no support from the doctor. I felt as though I was being viewed as a hypochondriac. I had a couple of phone consultations but never saw a doctor Eventually I was referred to a chest specialist for my cough she diagnosed gastric reflux. The second time I had Covid I was not so [ill] and I didn't consult my gp |
| 8 | **Care received: Yes, inferred as adequate**<br>**Perception of care experience: Positive**<br>**Ease of care access: Easy** | Intermediate, Female, 60-69 | Accessed out of hours for breathlessness. Seen very quickly and received excellent care, checked my pO2 and prescribed steroids. Breathlessness resolved. |
| 9 | Care received: Yes, inferred as adequate<br>Perception of care experience: Positive<br>Ease of care access: Easy<br>Received care from long COVID clinic or specialist | Relatively advantaged, Female, 50-59 | GP was very helpful, supportive and understanding when I had long covid in the first half of 2021. All over the phone of course but it still worked well. It was early days in terms of NHS support for long covid but the GP surgery tracked down the Yorkshire Rehabilitation Protocol (?) forms and submitted them into the system and I then had several phonecalls in the following few months from a long covid clinic. I had mostly recovered by then but it was really reassuring to know someone was there and it helped to speak to them. The app was launched not long after and also had good information in it. I was impressed at what the NHS provided, overall. |
| 10 | Care received: Yes, inferred as adequate<br>Perception of care experience: Positive<br>Ease of care access: Easy<br>Received care from long COVID clinic or specialist | Relatively advantaged, Male, 40-49 | I asked my GP for referral to long covid service and she did so after checking my symptoms. I saw a consultant and physio together (MDT). They gave me some advice and referred me to a novel treatment - hydrotherapy in a dead sea salt therapy pool. The pool sessions were useful, helpful & enjoyable. I was put onto PIFU and I re-contacted them when my symptoms flared up again. I self-referred to talking therapies. I have learned that long covid has significantly changed who I am - physically AND mentally. My former self image of a healthy, active, sporty man has been replaced by a tired, depressed and unwell person. This has had a significant impact on my mental health. |
| 11 | Care received: Yes, inferred as adequate<br>Perception of care experience: Positive<br>Ease of care access: Easy | Relatively disadvantaged, Female, 40-49 | Was able to speak to a nurse on the phone who happily prescribed whatever asthma meds and steroids to help my breathing - weren't testing generally then so couldn't tell if it was covid or not. Advice on 111 was very helpful eg pulseoximetry reading help and when to call 999 |
| 12 | Care received: Yes, inferred as adequate<br>Perception of care experience: Negative<br>Ease of care access: Difficult<br>Received care from long COVID clinic or specialist | Intermediate, Male, 50-59 | Long time (months) to see someone at a Long Covid clinic, clinic referred me on to other clinics to help rule other things out, but hard to contact or arrange an appointment with lots of different services. |
| 13 | Care received: Yes, perceived as inadequate<br>**Perception of care experience: Neutral or mixed**<br>Ease of care access: Difficult | Relatively disadvantaged, Female, 60-69 | Almost impossible to see the GP. I felt so poorly for weeks after. GP did arrange for a blood test and as that was 'normal' no further support available. My local pharmacist was amazing. Gave advice over the telephone whenever I called. |
| 14 | Care received: Yes, perceived as inadequate<br>Perception of care experience: Neutral or mixed<br>Ease of care access: Not stated<br>Received care from long COVID clinic or specialist | Intermediate, Female, 18-39 | GPs seemed to have limited knowledge of Long Covid condition, even 18 months into the pandemic. Switching to GP telephone triage appointment system was fine for me, and I could access my prescriptions online. Long Covid clinics have been helpful with more understanding and referrals and tests, but without a concrete condition to treat and discharge, I have reached the limit of what help they can offer me and am at a loss of what to do next. |
| 15 | Care received: Yes, inferred as adequate<br>Perception of care experience: Neutral or mixed<br>Ease of care access: Difficult | Relatively advantaged, Female, 70+ | It was hard to have contact with NHS GPs and Doctors. So I resorted to the private system using [PRIVATE PROVIDER NAME] and it was much better. A private GP helped me. |
| 16 | Care received: Yes, inferred as adequate<br>**Perception of care experience: Not clear**<br>Ease of care access: Difficult | Relatively disadvantaged, Female, 60-69 | I've been told had it not been for my tenacious attitude I wouldn't have had the many tests I have undergone. I have had three telephone consultations with a Respiratory Consultant and several telephone consultations with a respiratory physiotherapist and have now been referred to a Breathing Clinic where I am currently undergoing lung review tests etc. I was originally diagnosed with Bronchiectasis but this has now been thankfully downgraded to Long Covid. |

Just over half of respondents reported “excellent” or “very good” general health at the beginning of the COVID-19 pandemic (55%, Table S2). The proportion of respondents with a history of COVID-19 illness (83%), and prior or ongoing long COVID (22% with prior or ongoing self-reported long COVID, 9% with diagnosed long COVID, and 25% with ongoing symptoms at the time of the questionnaire), was higher than the general UK population at the time of questionnaire invitation, reflecting targeted recruitment into CSSB. The vast majority of self-reported COVID-19 illness was community based (8% of those with COVID-19 history accessed urgent care during illness), and confirmed by self-reported positive antibody or antigen tests (88% of those with COVID-19 history).

#### Associations between individual socio-demographic factors and health and social care access issues

The effects of individual pre-pandemic socio-demographic variables on health and social care access during the COVID-19 pandemic were estimated, using both linear regression to estimate the count of health and social care access issues (of 5) as a continuous outcome variable, and modified poisson regression to estimate the likelihood of one or more health care access (Figure 2, Table S2). Models controlled for potential confounding based on hypothesised DAG and used inverse probability weights to account for the probability of questionnaire completion.

**Figure 2.**
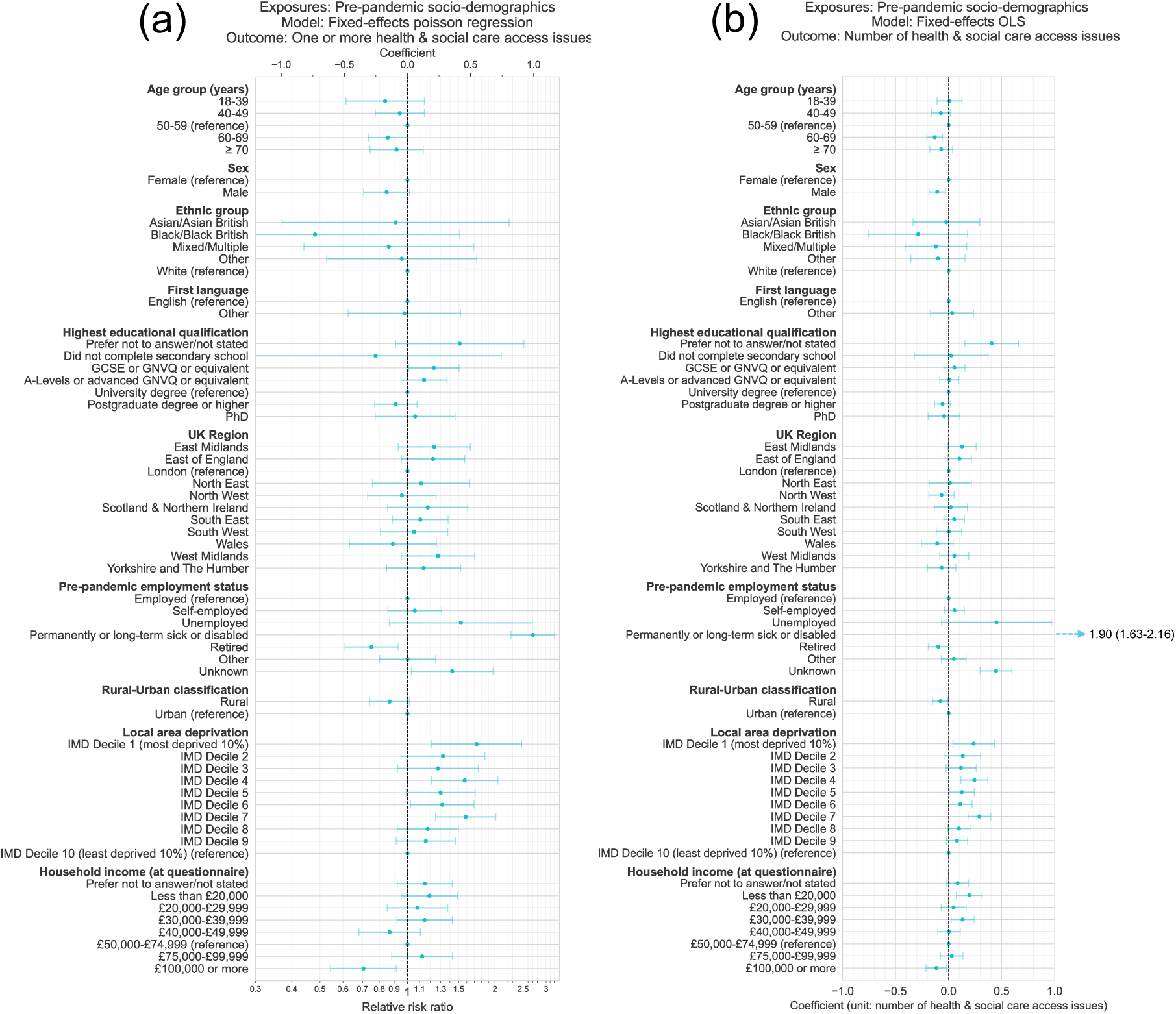
Coefficients and 95% confidence intervals from (a) poisson regression and (b) ordinary least squares linear regression models estimating associations between pre-pandemic socio-demographic factors and health & social care access issues during the COVID-19 pandemic. Results for each exposure variable originate from models with distinct adjustment variable sets to control for potential confounding based on hypothesised DAG as follows: Age: [Sex]; Sex: [Age]; Ethnic group: [Age]; First language: [Age, Sex, Ethnic group]; Education: [Age, Sex, Ethnic group, First language]; Region: [Age, Sex, Ethnic group, First language, Education]; Pre-pandemic employment status: [Age, Sex, Ethnic group, First language, Education, Region]; Rural-Urban classification (RUC): [Age, Sex, Ethnic group, First language, Education, Region, Pre-pandemic employment status]; Local area deprivation: [Age, Sex, Ethnic group, First language, Education, Region, Pre-pandemic employment status, RUC]; Household income (at time of questionnaire): [Age, Sex, Ethnicity, First language, Education, Region, Pre-pandemic employment status, RUC, Local area deprivation, Pre-pandemic health factors, COVID-19 illness factors]. Models included participant weighting for inverse probability of questionnaire completion.

Largest effects were observed for pre-pandemic employment status, where those long term sick or disabled reported a much higher average number of care access issues vs. employed participants (OLS coefficient: 1.90, 95% CI: 1.63-2.16); poisson regression RR: 2.70, 95% CI: 2.27-3.21).

Effects were also observed in both linear and poisson regression models for those living in more deprived areas, who were more likely to report issues vs. least deprived 10% of areas (most deprived 10%, OLS: 0.24, 0.04-0.43; RR: 1.73, 1.21-2.47). Fewer issues were also observed for those with household income of more than £100,000 at the time of the questionnaire vs. £50,000-74,999 (OLS: - 0.12, -0.21 - -0.02; RR: 0.71, 0.54-0.92), and for those aged 60-69 vs. 50-59 (with no other clear trend with age) (OLS: -0.13, -0.20 - -0.06; RR: 0.86, 0.73-1.00).

Lower number of access issues were also observed, but in linear regression models only, and with smaller effect sizes, for male vs. female participants, those living in rural vs. urban areas, for those retired vs. employed at the start of the COVID-19 pandemic, and for those with undergraduate degree level vs. those who preferred not to disclose their education level. Finally, increased likelihood of access issues was also observed, but in poisson regression models only, for participants with GCSE or GNVQ or equivalent (typically finishing education at age 16) vs. undergraduate degree level.

#### Intersectional MAIHDA analysis of inequalities in health & social care access issues

In addition to estimating associations between individual pre-pandemic socio-demographic variables and health and social care access during the COVID-19 pandemic, intersectional MAIHDA analysis was performed to estimate associations between ‘social strata’, with particular combinations of sex, education level and local area deprivation, and care access, while taking into account interaction effects (Figure 3, Table S3, Table S4).

**Figure 3.**
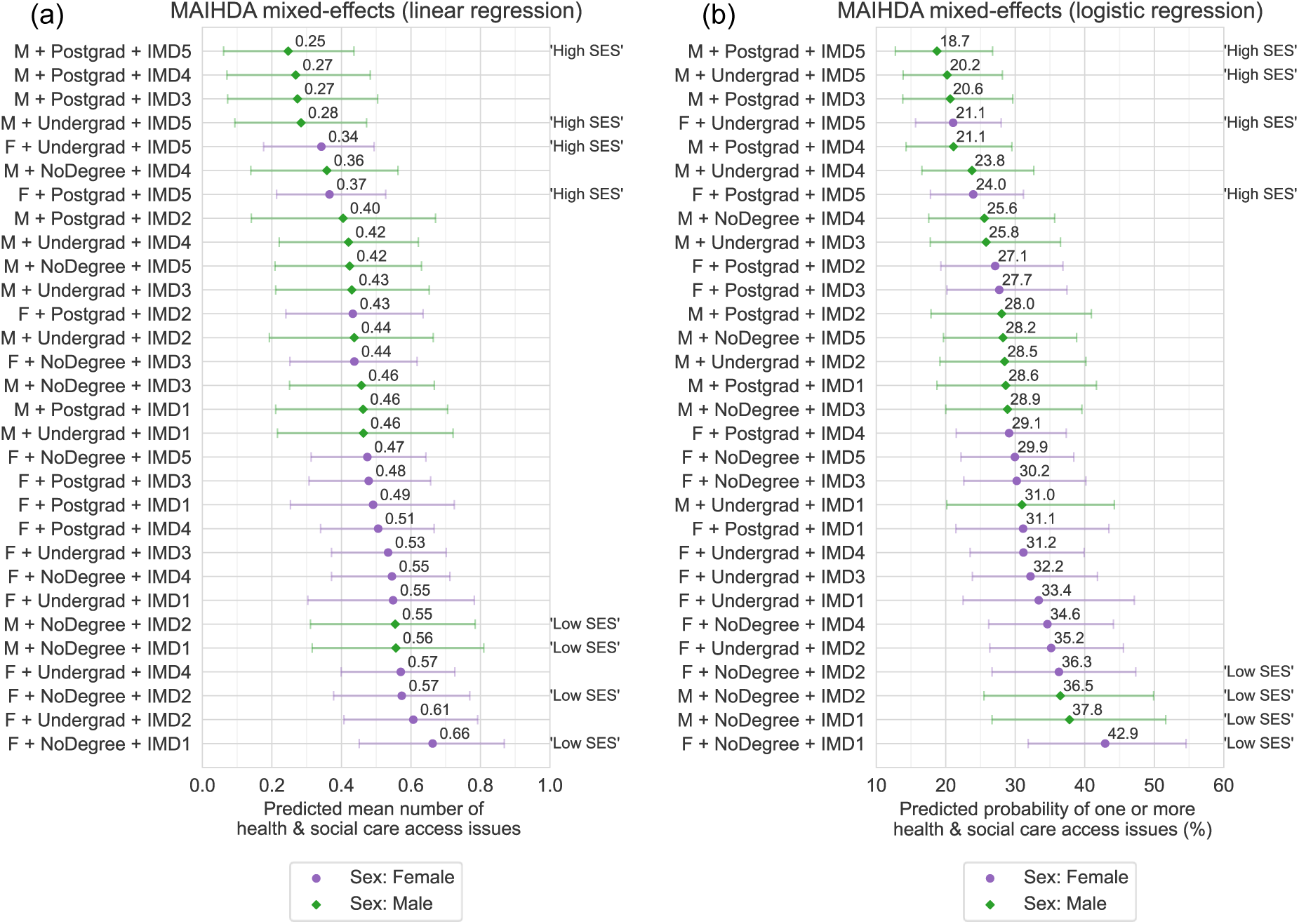
Intersectional MAIHDA models estimating effect of combinations of sex, education level and local area deprivation on health & social care access issues during the COVID-19 pandemic. (a) Predicted mean (with approximate 95% confidence intervals) number of issues from linear regression and (b) predicted probability (with approximate 95% confidence intervals) of one or more issues from logistic regression. Fixed effects included in models were age group, ethnic group, sex, education level and local area deprivation. Models included participant weighting for inverse probability of questionnaire completion. Strata labels: F = Female, M = Male; NoDegree = Less than degree level education (including not stated/prefer not to say), Undergrad = Undergraduate degree level, Postgrad = Postgraduate degree level or higher; IMD1-IMD5 = Index of Multiple Deprivation Quintile 1 to 5, where 1 is most deprived 20% of areas, and 5 is least deprived 20%. Labels “High SES” are given to highlight strata where the combination of education level and deprivation suggest relatively high socio-economic status - living in the least deprived quintile 1 and having a degree level education or higher. “Low SES” highlights relatively low socio-economic status strata – living in the most deprived 40% (quintile 4 or 5) and having less than degree level education.

MAIHDA intersectional models found large variation in health & social care issues between strata, with the predicted mean number of issues varying between 0.25 (approximate 95% CI: 0.06-0.44) and 0.66 (0.45-0.87), and likelihood of one or more varying between 18.7% (12.8%-26.7%) and 42.9% (31.9%-54.6%). Female sex strata tended to have a higher probability of having one or more health and social care issues, and a higher predicted mean number of issues, than male strata. However, male as well as female sex strata with socio-economically disadvantageous combinations of both education and local area deprivation – i.e. low education levels and living in high deprivation areas (‘Low socio-economic status (SES)’ strata) – were consistently the most likely to report health and social care access issues. In contrast, both female and male sex strata with high education levels living in low deprivation areas (‘High SES’) were among those least likely to report issues.

The variance partition coefficients (VPC) of MAIHDA models were low (VPC = 0.96% for both linear and logistic regression models), indicating that the vast majority of the variation in care access issues between strata are accounted for by additive effects of individual variables, with interaction effects making a relatively small contribution.

In supplementary analysis, additional MAIHDA models were run on samples stratified by COVID-19 history (Figure S3). The size of the inequality in care access issues between strata was slightly larger for the ‘no COVID’ and ‘long COVID’ subsamples, compared to the ‘short COVID’ subsample.

Similar to the overall sample, the likelihood of care access issues was higher for female strata and ordered along the lines of socio-economic status in all stratified subsamples, albeit with small differences in the relative importance of deprivation and education.

To further evaluate the role of economic capital on access to care, household income was also included in the social strata variable (Figure S4). It is noted that data represented income at the time of the questionnaire, and so may be subject to reverse causality if health care access issues during the pandemic caused declines in income. Nevertheless, after including income, strata with the highest predicted probability of health and social care access issues were consistently those with the lowest income band (< £20,000), in combination with fewer educational qualifications and/or living in higher deprivation areas, while strata with lowest predicted probability were those with the highest income band (≥ £100,000) in combination with higher education level and/or living in lower deprivation areas. The range of the inequality across strata was also increased with addition of income for both linear regression (range = 0.71 vs. 0.41) and logistic (range = 34% vs 24%) models.

In final sensitivity analyses, negligible changes were observed in MAIHDA model predictions after additional adjustment for pre-pandemic health factors (themselves tested for associations, with results presented in Table S2), representing an alternative hypothesised causal diagram where health precedes social factors (vs. social factors preceding health factors that is assumed in the main analysis) (Figure S5).

### Part 2: Health care experiences for COVID-19 (open question analysis)

#### COVID-19 health care experiences from qualitative coding

To explore health care experiences specifically for COVID-19 illness in more detail, free-text responses to an open survey question on health care experiences for COVID-19 were coded using a deductive approach for a subset of respondents (N = 335) (outcomes given in Table 1, full codes and frequencies given in Table S5). The subset was comprised of three approximately equal sized groups selected to assess experiences of respondents with varying degrees of advantaged or disadvantaged positions/statuses/characteristics (selection criteria are given in Table 2, Table 3, socio-demographic characteristics of the subset are given in Table S6).

Quantitative analysis of qualitatively coded open text responses found socio-demographic gradients across multiple aspects of COVID-19 health care experiences (Figure 4, with illustrative quotes given in Table 5). Despite the small sample size of the qualitatively coded subset, multiple observations of poorer care access and experiences for more disadvantaged vs. more advantaged participant groups were statistically significant when tested in multivariable poisson regression models adjusting for age group (Figure S6). These observations were: lower likelihood of feeling that they have received adequate care, among those who expressed need for care (RR = 0.69, 95% CI: 0.50-0.96, p = 0.03 for the relatively disadvantaged group vs. the relatively advantaged group from multivariable regression models adjusting for age group); lower likelihood of receiving specialist care for long COVID among those with a long COVID diagnosis (RR = 0.14, 95% CI: 0.04-0.50, p = 0.003 for relatively disadvantaged vs. relatively advantaged); and higher likelihood of negative experiences (RR = 2.44, 95% CI: 1.24-4.80, p = 0.01 for relatively disadvantaged vs. relatively advantaged). Also of note, but not significant at a p < 0.05 level, were apparent higher proportions of relatively disadvantaged participants reporting unsuccessful attempts to obtain care, and easier access to care among relatively advantaged participants.

**Figure 4.**
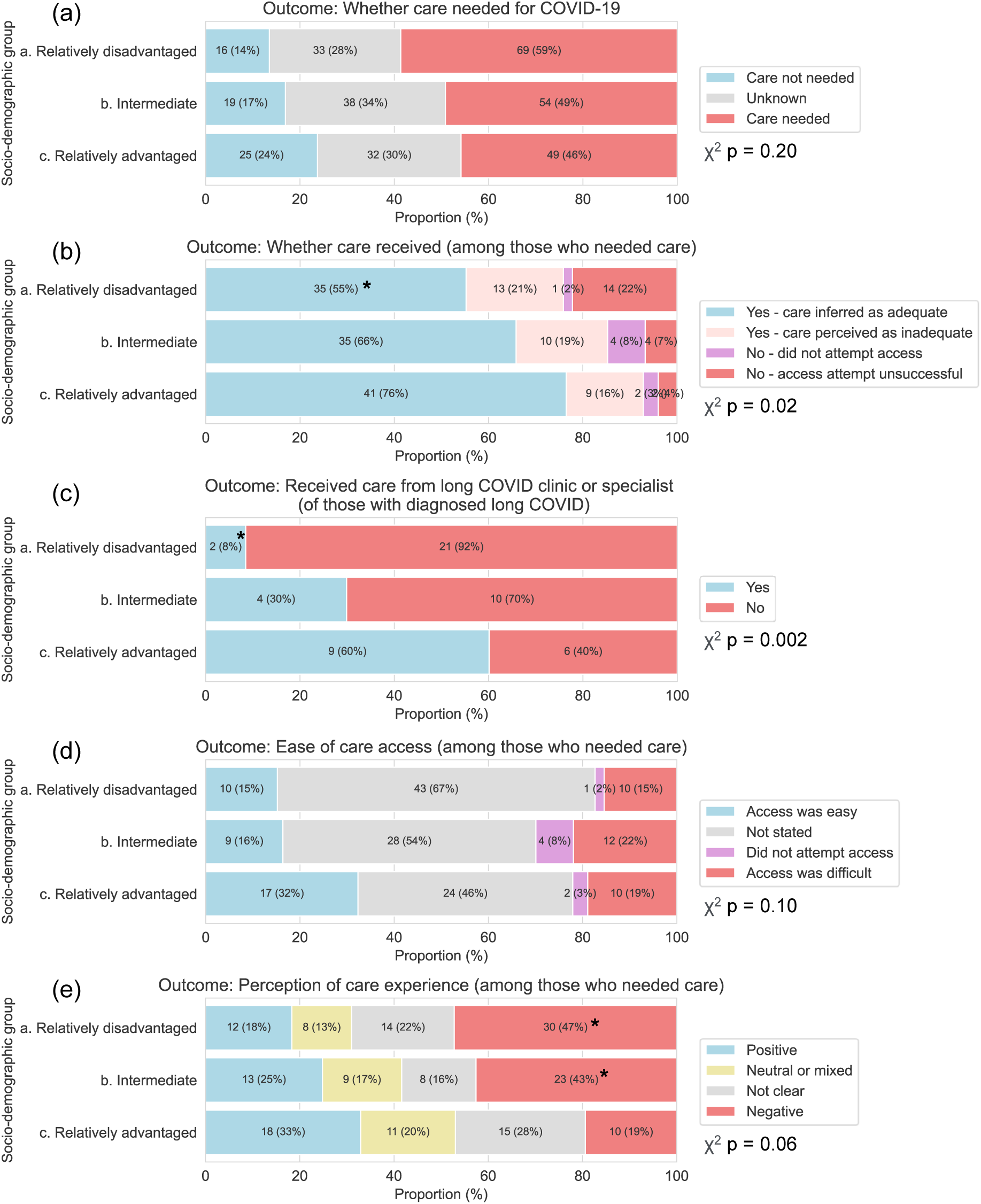
Frequency of various COVID-19 health care experiences from qualitative coding of free-text responses. Bar labels present pseudo-population sample sizes and proportions after weighting for the inverse probability of selection into the qualitatively coded subset (a) and needing care for COVID-19 (b to e). Bars with * highlight where differences in comparison with relatively advantaged group were statistically significant, p < 0.05, from multivariable poisson regression models adjusting for age group (presented in Figure S6). P-values for chi-squared tests of independence are also presented.

The contrasts in experiences between participants with different socio-economic positions are exemplified by comparing illustrative quotes 9 and 5 from Table 5:

“GP was very helpful, supportive and understanding when I had long covid in the first half of 2021. All over the phone of course but it still worked well. It was early days in terms of NHS support for long covid but the GP surgery tracked down the Yorkshire Rehabilitation Protocol (?) forms and submitted them into the system and I then had several phonecalls in the following few months from a long covid clinic. I had mostly recovered by then but it was really reassuring to know someone was there and it helped to speak to them. The app was launched not long after and also had good information in it. I was impressed at what the NHS provided, overall.” **Quote 9. Relatively advantaged, Female, 50-59.**

“It was really hard to see GP and still is. I am still waiting for specialist lung appointment and I [have] a nodule on my lung, my cough is awful and I am always tired. The health care is rubbish since covid.” **Quote 5. Relatively disadvantaged, Female, 50-59.**

Both quotes were made by female participants of white ethnic groups in their 50s, both living in the same region of England, and both with diagnoses of long COVID. However, the participant who wrote quote 9 was part of the relatively advantaged group, by virtue of having postgraduate level of education, being in the highest income bracket (≥ £100,000), and living in an area among the least deprived 30% in the UK. In contrast, quote 5’s participant was part of the relatively disadvantaged group, with a less than degree level of education, below median level of household income (< £40,000), and lived in an area among the most deprived 30% in the UK.

Unsuccessful attempts to access care and negative experiences most often concerned not being able to get appointments with doctors in primary care. Several responses also described having long waiting times for long COVID clinics, and difficulties obtaining relevant treatment from GPs (e.g., Table 5, quote 4, 5 & 6). Care ascribed as inadequate often described feeling unsupported by health professionals and having a lack of treatment options (e.g., Table 5, quote 6, 7 & 14). In contrast, positive experiences often described fast and easy access to care, referrals to specialist services, and receipt of often multiple treatments (e.g., Table 5, quote 10). Almost all positive experiences among relatively advantaged participants mentioned their GP (e.g., Table 5, quote 10), while relatively disadvantaged participants mentioned using other services such as pharmacy and 111 or 999 more frequently (e.g., Table 5, quote 11).

#### Illustrative quotes

## Discussion

### Key points

Analysis of both closed and open survey questions emphasised the cumulative effects of holding multiple socially advantaged or disadvantaged positions on health care access and experiences among participants of the COVID Symptom Study Biobank UK longitudinal population study.

In the first part of our study, analysis of a closed survey question from participants of the COVID Symptom Study Biobank UK longitudinal population study found large socio-demographic inequalities in the frequency of issues accessing health and social care in the first two and a half years of the COVID-19 pandemic in the UK. Gradients were found along the lines of social power and socio-economic status when looking at variation based on explicit combinations of sex, education, local area deprivation and income, with more advantaged participants less likely to experience issues accessing care (Figure 3). Female strata consistently reported more care access issues, while the highest and lowest probabilities of issues were seen for the most socio-economically advantaged or disadvantaged strata among both females and males. Associations were not explained by pre-pandemic health (Figure S5), and similar socio-economic gradients were also observed within groups with comparable histories of COVID-19 illness (Figure S3), suggesting that social inequalities in access to care exist independent of the specific illnesses for which care is needed.

Similar social gradients were then observed in quantitative analysis of a subset of deductively coded free-text responses to an open question about health care experiences for COVID-19 illness (Figure 4). Analysis suggested less frequent access to adequate care and higher frequency of negative experiences for more disadvantaged participants. Receipt of long COVID specialist care was also socially patterned, even among those who had a long COVID diagnosis. Multiple observations were statistically significant despite the subset’s small sample size. Illustrative quotes provided detailed accounts of the multiple aspects of care that were found to vary with social circumstances (Table 5).

### Interpretation

Observed inequalities in health care may be explained both in terms of structural differences at an area level, and individual level differences in social, cultural, and economic capitals, following Bourdieu’s conceptualisation of social class [51], as well as the related concept of health capital [52].

At a structural level, lower funding and workforce provision in primary health care services in more deprived areas [53,54] are likely to contribute to inequalities we observe, perhaps most significantly to initial access to care and waiting times, as well as poorer interpersonal experiences due to greater time constraints. While higher personal income may provide the financial means to access care privately, differences in other forms of individual capital may explain more of the observed inequalities in care experiences beyond initial access. Higher cultural or health capital, such as greater education or understanding of health and health care services, may help more advantaged patients to self-advocate at a time where persistence appears to be key [25]. Meanwhile, social capital, in the form of knowing health care professionals, may aid navigation through a complex and fragmented health care system. Stark contrasts in interpersonal experiences with health care professionals (mostly GPs) between advantaged and disadvantaged participants also echo well-known social gradients in patient-doctor communication favouring higher socio-economic status patients [55], and may be explained in part by bias from health care providers against lower socio-economic status patients, which appears to negatively affect clinical decision making [56].

Considering health care specifically for long COVID, the poorer access to both general and specialist long COVID health care for less advantaged participants is further evidence of an inverse care law in action. Less frequent receipt of specialist care despite having a long COVID diagnosis was particularly concerning (Figure 4 (c)). Furthermore, the repeated description by participants living with long COVID of feeling like they had no treatment options emphasises the need for continued support for patients as well as the need for further research to build upon recent trials of rehabilitation programs [34], and improve efficacy of UK post-COVID clinics [57].

### Limitations

We note the limitations in our study. Recruitment into the CSSB was conditioned on the use of a smartphone app and targeted based on COVID-19 histories. As such, the cohort over-represents experiences of people with history of long COVID, as well as female sex, white ethnic groups and more socio-economically advantaged groups compared to the UK population, and so results may not be generalisable to the UK as a whole.

Our analysis samples were subject to CSSB members voluntary participation in surveys, as well as selection of subsets, which may be subject to selection bias. We attempted to mitigate bias by using inverse probability weighting, with a dedicated set of weights generated for each analysis subset.

Choice of factors to examine in intersectional MAIHDA models was limited by sample size, for example inclusion of ethnic group was not feasible due to small numbers of racially minoritised ethnic groups. Similarly, the number of open-text responses able to be coded by hand was limited, and so we were not able to analyse explicit intersectional inequalities in COVID-19 using an approach such as MAIHDA.

### Summary

In summary, our study suggests there were large intersectional inequalities in health care access and experiences during the COVID-19 pandemic in the UK, for people with or without history of COVID-19 illness, along the lines of structural societal advantage. Further research could focus on identifying how much of observed inequalities in health care quality arise due to differences in interpersonal vs. structural factors, in order to address issues in both initial access to health care services and experiences upon entry. Finally, further evidence of greater unmet need for care among disadvantaged participants living with long COVID emphasises the continued need for support that works for those with least power and status within society.

## Contributors

Following CRediT framework: https://www.elsevier.com/authors/policies-and-guidelines/credit-author-statement.

Conceptualisation: NJC, CJS, with contribution from all other authors (AB, ABC, VB, JDC, The COVID Symptom Study Biobank Consortium)

Funding acquisition: CJS Project administration: VB, CJS

Methodology: NJC, CJS, JDC, AB, ABC Formal analysis: NJC, AB, ABC Investigation: VB, NJC

Visualisation: NJC

Data curation: VB, NJC, AB, ABC

Writing – original draft: NJC

Writing – review and editing: NJC, CJS, AB, ABC, VB, JDC, The COVID Symptom Study Biobank Consortium

NJC accepts full responsibility for the finished work and/or the conduct of the study, had access to the data, and controlled the decision to publish.

## Data sharing statement

For the purposes of open access, the author has applied a Creative Commons Attribution (CC BY) licence to any Accepted Author Manuscript version arising from this submission.

Access to data in the COVID Symptom Study Biobank is available to bona fide health researchers on application to the COVID Symptom Study Biobank Management Group. Further details are available online at https://cssbiobank.com/information-for-researchers including application forms and contact information.

Analysis code used in this study is available openly on GitHub at https://github.com/nathan-cheetham/CSSBiobank_CareExperiences (following peer review). Anonymised COVID Symptom Study data are available to researchers to be shared with researchers according to their protocols in the public interest through Health Data Research UK (HDRUK) and Secure Anonymised Information Linkage consortium, housed in the UK Secure Research Platform (Swansea, UK) at https://web.www.healthdatagateway.org/dataset/fddcb382-3051-4394-8436-b92295f14259.

## Ethics statement

Yorkshire & Humber NHS Research Ethics Committee gave ethical approval for the COVID Symptom Study Biobank, Ref: 20/YH/0298.

## Supporting information

STROBE checklist

## Data Availability

For the purposes of open access, the author has applied a Creative Commons Attribution (CC BY) licence to any Accepted Author Manuscript version arising from this submission.
Access to data in the COVID Symptom Study Biobank is available to bona fide health researchers on application to the COVID Symptom Study Biobank Management Group. Further details are available online at https://cssbiobank.com/information-for-researchers including application forms and contact information.
Analysis code used in this study is available openly on GitHub at https://github.com/nathan-cheetham/CSSBiobank_CareExperiences (following peer review). Anonymised COVID Symptom Study data are available to researchers to be shared with researchers according to their protocols in the public interest through Health Data Research UK (HDRUK) and Secure Anonymised Information Linkage consortium, housed in the UK Secure Research Platform (Swansea, UK) at https://web.www.healthdatagateway.org/dataset/fddcb382-3051-4394-8436-b92295f14259.

## Acknowledgements

We thank COVID Symptom Study Biobank participants, in particular those who participated in various studies while experiencing ongoing symptoms. We are grateful to the CSS Biobank Volunteer Advisory Panel for their input on the development of the biobank, and feedback on the design of this study and its significance for patients and the wider public.

We are grateful to staff at Zoe Ltd (including Christina Hu and Joan Capdevila Pujol) for their work on the CSS app, for enabling recruitment to the CSS Biobank and sharing and maintaining CSS data. We thank Katie Doores, Michael Malim, Carl Graham, Jeffrey Seow, Sam Acors, and Neo Kouphou for antibody testing of participant blood samples.

## References

1 Maddock J, Parsons S, Di Gessa G, et al. Inequalities in healthcare disruptions during the COVID-19 pandemic: evidence from 12 UK population-based longitudinal studies. BMJ Open 2022;12:e064981. doi:10.1136/BMJOPEN-2022-064981

2 Topriceanu CC, Wong A, Moon JC, et al. Evaluating access to health and care services during lockdown by the COVID-19 survey in five UK national longitudinal studies. BMJ Open 2021;11:e045813. doi:10.1136/BMJOPEN-2020-045813

3 Cheetham NJ, Bowyer V, García MP, et al. Social determinants of recovery from ongoing symptoms following COVID-19 in two UK longitudinal studies: a prospective cohort study. medRxiv 2023;:A59.1–A59. doi:10.1101/2023.12.21.23300125

4 Centre for Ageing Better. Ethnic health inequalities in later life: The persistence of disadvantage from 1993-2017 (Centre for Ageing Better). 2021.

5 Wu Y-T, Daskalopoulou C, Muniz Terrera G, et al. Education and wealth inequalities in healthy ageing in eight harmonised cohorts in the ATHLOS consortium: a population-based study. The Lancet Public Health 2020;5:e386–94. doi:10.1016/S2468-2667(20)30077-3

6 Parker M, Bucknall M, Jagger C, et al. Population-based estimates of healthy working life expectancy in England at age 50 years: analysis of data from the English Longitudinal Study of Ageing. The Lancet Public Health 2020;5:e395–403. doi:10.1016/S2468-2667(20)30114-6

7 Trend in life expectancy by National Statistics Socioeconomic Classification, England and Wales: 1982 to 1986 and 2012 to 2016 - Office for National Statistics. 2022.https://www.ons.gov.uk/peoplepopulationandcommunity/birthsdeathsandmarriages/lifeexpectancies/bulletins/trendinlifeexpectancyatbirthandatage65bysocioeconomicpositionbasedonthenationalstatisticssocioeconomicclassificationenglandandwales/1982to1986and2012to (accessed 13 Jan 2025).

8 Metsis K, Inchley J, Williams AJ, et al. What can the 2001, 2011 and 2021 UK censuses tell us about health inequalities among young people? A cross-sectional study using censuses from England, Wales, and Scotland. Research Square 2024.

9 The Health Foundation. Relationship between income and health | The Health Foundation. 2024.https://www.health.org.uk/evidence-hub/money-and-resources/income/relationship-between-income-and-health (accessed 13 Jan 2025).

10 Office for National Statistics. General health by age, sex and deprivation, England and Wales: Census 2021 - Office for National Statistics. 2023. https://www.ons.gov.uk/peoplepopulationandcommunity/healthandsocialcare/healthandwellbeing/datasets/generalhealthbyagesexanddeprivationenglandandwales (accessed 11 Dec 2023).

11 PHE. Disparities in the risk and outcomes of COVID-19. 2020. https://www.gov.uk/government/publications/covid-19-review-of-disparities-in-risks-and-outcomes (accessed 17 Nov 2021).

12 Barnard S, Fryers P, Fitzpatrick J, et al. Inequalities in excess premature mortality in England during the COVID-19 pandemic: a cross-sectional analysis of cumulative excess mortality by area deprivation and ethnicity. BMJ Open 2021;11:e052646. doi:10.1136/BMJOPEN-2021-052646

13 Hastie CE, Lowe DJ, McAuley A, et al. Outcomes among confirmed cases and a matched comparison group in the Long-COVID in Scotland study. Nature Communications 2022;13:5663. doi:10.1038/s41467-022-33415-5

14 Office for National Statistics. Self-reported coronavirus (COVID-19) infections and associated symptoms, England and Scotland: November 2023 to March 2024. https://www.ons.gov.uk/peoplepopulationandcommunity/healthandsocialcare/conditionsanddiseases/articles/selfreportedcoronaviruscovid19infectionsandassociatedsymptomsenglandandscotland/november2023tomarch2024 (accessed 6 May 2025).

15 Shabnam S, Razieh C, Dambha-Miller H, et al. Socioeconomic inequalities of Long COVID: a retrospective population-based cohort study in the United Kingdom. Journal of the Royal Society of Medicine 2023;116:263–73. doi:10.1177/01410768231168377

16 Greenhalgh T, Sivan M, Perlowski A, et al. Long COVID: a clinical update. The Lancet 2024;404:707–24. doi:10.1016/S0140-6736(24)01136-X

17 ONS. Prevalence of ongoing symptoms following coronavirus (COVID-19) infection in the UK: 30 March 2023 - Office for National Statistics. 2023.https://www.ons.gov.uk/peoplepopulationandcommunity/healthandsocialcare/conditionsanddiseases/bulletins/prevalenceofongoingsymptomsfollowingcoronaviruscovid19infectionintheuk/30march2023 (accessed 25 Apr 2023).

18 Wang H-I, Doran T, Crooks MG, et al. Prevalence, risk factors and characterisation of individuals with long COVID using Electronic Health Records in over 1.5 million COVID cases in England. Journal of Infection 2024;89:106235. doi:10.1016/j.jinf.2024.106235

19 Knuppel A, Boyd A, Macleod J, et al. The long COVID evidence gap in England. The Lancet 2024;403:1981–2. doi:10.1016/S0140-6736(24)00744-X

20 Henderson AD, Butler-Cole BF, Tazare J, et al. Clinical coding of long COVID in primary care 2020–2023 in a cohort of 19 million adults: an OpenSAFELY analysis. eClinicalMedicine 2024;72:102638. doi:10.1016/j.eclinm.2024.102638

21 Sunkersing D, Goodfellow H, Mu Y, et al. Long COVID symptoms and demographic associations: A retrospective case series study using healthcare application data. JRSM Open 2024;15. doi:10.1177/20542704241274292

22 Tudor Hart J. The inverse care law. The Lancet 1971;297:405–12. doi:10.1016/S0140-6736(71)92410-X

23 Baz SA, Fang C, Carpentieri JD, et al. ‘I don’t know what to do or where to go’. Experiences of accessing healthcare support from the perspectives of people living with Long Covid and healthcare professionals: A qualitative study in Bradford, UK. Health Expectations 2023;26:542–54. doi:10.1111/hex.13687

24 Duncan E, Alexander L, Cowie J, et al. Investigating Scottish Long COVID community rehabilitation service models from the perspectives of people living with Long COVID and healthcare professionals: a qualitative descriptive study. BMJ Open 2023;13:e078740. doi:10.1136/bmjopen-2023-078740

25 Fang C, Baz SA, Sheard L, et al. “They seemed to be like cogs working in different directions”: a longitudinal qualitative study on Long COVID healthcare services in the United Kingdom from a person-centred lens. BMC Health Services Research 2024;24:406. doi:10.1186/s12913-024-10891-7

26 Clutterbuck D, Ramasawmy M, Pantelic M, et al. Barriers to healthcare access and experiences of stigma: Findings from a coproduced Long Covid case-finding study. Health Expectations 2024;27. doi:10.1111/hex.14037

27 Gamillscheg P, Łaszewska A, Kirchner S, et al. Barriers and facilitators of healthcare access for long COVID-19 patients in a universal healthcare system: qualitative evidence from Austria. International Journal for Equity in Health 2024;23:220. doi:10.1186/s12939-024-02302-4

28 Turk F, Sweetman J, Chew-Graham CA, et al. Accessing care for Long Covid from the perspectives of patients and healthcare practitioners: A qualitative study. Health Expectations 2024;27:e14008. doi:10.1111/hex.14008

29 Smyth N, Ridge D, Kingstone T, et al. People from ethnic minorities seeking help for long COVID: a qualitative study. British Journal of General Practice 2024;74:e814–22. doi:10.3399/BJGP.2023.0631

30 Yong A, Germain S. Ethnic minority and migrant women’s struggles in accessing healthcare during COVID-19: an intersectional analysis. Journal for Cultural Research 2022;26:65–82. doi:10.1080/14797585.2021.2012090

31 Baz SA, Woodrow M, Clutterbuck D, et al. Long COVID and Health Inequalities: What’s Next for Research and Policy Advocacy? Health Expectations 2024;27:e70047. doi:10.1111/HEX.70047

32 Davis HE, McCorkell L, Vogel JM, et al. Long COVID: major findings, mechanisms and recommendations. Nature Reviews Microbiology 2023;:1–14. doi:10.1038/s41579-022-00846-2

33 Bramante CT, Buse JB, Liebovitz DM, et al. Outpatient treatment of COVID-19 and incidence of post-COVID-19 condition over 10 months (COVID-OUT): a multicentre, randomised, quadruple-blind, parallel-group, phase 3 trial. The Lancet Infectious Diseases 2023;0. doi:10.1016/S1473-3099(23)00299-2

34 Nerli TF, Selvakumar J, Cvejic E, et al. Brief Outpatient Rehabilitation Program for Post– COVID-19 Condition: A Randomized Clinical Trial. JAMA Network Open 2024;7:e2450744–e2450744. doi:10.1001/JAMANETWORKOPEN.2024.50744

35 Love HR, Corr C. Integrating Without Quantitizing: Two Examples of Deductive Analysis Strategies Within Qualitatively Driven Mixed Methods Research. Journal of Mixed Methods Research 2022;16:64–87. doi:10.1177/1558689821989833

36 Miles MB., Huberman AM., Saldaña J. Qualitative data analysis : a methods sourcebook. SAGE Publications, Inc. 2014.

37 Campbell JL, Quincy C, Osserman J, et al. Coding In-depth Semistructured Interviews: Problems of Unitization and Intercoder Reliability and Agreement. Sociological Methods & Research 2013;42:294–320. doi:10.1177/0049124113500475

38 Jones CP. Systems of power, axes of inequity: Parallels, intersections, braiding the strands. Medical Care 2014;52:S71–5. doi:10.1097/MLR.0000000000000216

39 Zou G. A Modified Poisson Regression Approach to Prospective Studies with Binary Data. American Journal of Epidemiology 2004;159:702–6. doi:10.1093/AJE/KWH090

40 Mansournia MA, Nazemipour M, Naimi AI, et al. Reflection on modern methods: demystifying robust standard errors for epidemiologists. International Journal of Epidemiology 2021;50:346–51. doi:10.1093/IJE/DYAA260

41 Textor J, van der Zander B, Gilthorpe MS, et al. Robust causal inference using directed acyclic graphs: the R package ‘dagitty’. International Journal of Epidemiology 2017;45:dyw341. doi:10.1093/ije/dyw341

42 Evans CR, Leckie G, Subramanian S V., et al. A tutorial for conducting intersectional multilevel analysis of individual heterogeneity and discriminatory accuracy (MAIHDA). SSM - Population Health 2024;26:101664. doi:10.1016/J.SSMPH.2024.101664

43 Evans CR, Borrell LN, Bell A, et al. Clarifications on the intersectional MAIHDA approach: A conceptual guide and response to Wilkes and Karimi (2024). Social Science & Medicine 2024;350:116898. doi:10.1016/J.SOCSCIMED.2024.116898

44 Crenshaw K. Mapping the Margins: Intersectionality, Identity Politics, and Violence against Women of Color. Stanford Law Review 1991;43:1241. doi:10.2307/1229039

45 Collins PH. Intersectionality’s Definitional Dilemmas. Annual Review of Sociology 2015;41:1–20. doi:10.1146/annurev-soc-073014-112142

46 Bell A. OSF | MAIHDA Tutorial Code.R. 2024. doi:10.17605/OSF.IO/DTVC3

47 Fernández-Sanlés A, Smith D, Clayton GL, et al. Bias from questionnaire invitation and response in COVID-19 research: an example using ALSPAC. Wellcome Open Research 2021;6:184. doi:10.12688/wellcomeopenres.17041.1

48 Griffith GJ, Morris TT, Tudball MJ, et al. Collider bias undermines our understanding of COVID-19 disease risk and severity. Nature Communications 2020;11:1–12. doi:10.1038/s41467-020-19478-2

49 van Zwieten A, Tennant PWG, Kelly-Irving M, et al. Avoiding overadjustment bias in social epidemiology through appropriate covariate selection: a primer. Journal of Clinical Epidemiology 2022;149:127–36. doi:10.1016/j.jclinepi.2022.05.021

50 Cheetham NJ, Penfold R, Giunchiglia V, et al. The effects of COVID-19 on cognitive performance in a community-based cohort: a COVID symptom study biobank prospective cohort study. eClinicalMedicine 2023;62:102086. doi:10.1016/j.eclinm.2023.102086

51 Bourdieu P. Distinction: A Social Critique of the Judgement of Taste. Harvard University Press 1984.

52 Schneider-Kamp A. Health capital: toward a conceptual framework for understanding the construction of individual health. Social Theory & Health 2021;19:205–19. doi:10.1057/s41285-020-00145-x

53 Fisher R, Dunn P, Asaria M, et al. Level or not? Comparing general practice in areas of high and low socioeconomic deprivation in England. 2020. https://www.health.org.uk/publications/reports/level-or-not

54 Nussbaum C, Massou E, Fisher R, et al. Inequalities in the distribution of the general practice workforce in England: a practice-level longitudinal analysis. BJGP Open 2021;5:1–10. doi:10.3399/BJGPO.2021.0066

55 Verlinde E, De Laender N, De Maesschalck S, et al. The social gradient in doctor-patient communication. International Journal for Equity in Health 2012;11:12. doi:10.1186/1475-9276-11-12

56 Job C, Adenipekun B, Cleves A, et al. Health professionals implicit bias of patients with low socioeconomic status (SES) and its effects on clinical decision-making: a scoping review. BMJ Open 2024;14:e081723. doi:10.1136/bmjopen-2023-081723

57 Roman M, Lawrence R, Osborne T, et al. A National Evaluation of Outcomes in Long COVID Services using Digital PROM Data from the ELAROS Platform. 2023. https://locomotion.leeds.ac.uk/wp-content/uploads/sites/74/2023/10/National-Evaluation-of-LC-Service-Outcomes-using-ELAROS-Data-09-10-23.pdf

