## Supplementary information for "Socio-demographic inequalities in COVID-19 health care access and experiences in the United Kingdom: intersectional and mixed-methods analyses of open and closed questions in a prospective cohort study"

#### Authors & affiliations

Nathan J. Cheetham[1]*, Anoushka Beattie[2], Alastair B. Comery[3], Vicky Bowyer[1], The COVID Symptom Study Biobank Consortium, J. D. Carpentieri[2], Claire J. Steves[1,4]*

* Corresponding authors

1 Department of Twin Research and Genetic Epidemiology, King's College London, London, United Kingdom

2 Institute of Education, University College London, London, United Kingdom

3 Centre for Death and Society, Department of Social Policy Sciences, University of Bath, Bath, United Kingdom

4 Guy's & St Thomas's NHS Foundation Trust, London, United Kingdom

#### Section S1 Full variable details

*Socio-demographic characteristics*

Information on age group at the time of the 2022 CSSB questionnaire was derived from date of birth self-reported at CSSB consent (2022 – Year of Birth).

Information on sex assigned at birth (female/intersex/male/prefer not to say) was self-reported at registration to the CSS app.

First language was self-reported in the August, 2022 CSSB questionnaire, and grouped into English and Other due to small number of individuals with first languages other than English.

Information on race/ethnicity was collected at CSSB consent with the question "what is your ethnic group?", using UK 2021 census categories (Asian/Asian British, Black/Black British/Caribbean/African, Mixed/Multiple ethnic groups, White, and Other ethnic groups) [1].

Information on highest educational qualification was collected in the August, 2022 CSSB questionnaire "What is the highest academic/educational qualification (or its nearest equivalent) you have received?".

Local area deprivation, UK geographic region of residence and Rural-Urban classification (most recent version from each UK nation) were derived from address data collected upon joining the CSS app and CSSB consent (with precedence given to earlier CSS app data). Deprivation was measured by the Index of Multiple Deprivation (IMD) quantiles of lower super output area rank, with data from England 2019 [2], Wales 2019 [3], Scotland 2020 [4], and Northern Ireland 2017 [5]. Individuals living in Scotland and Northern Ireland were grouped together in a "Scotland/Northern Ireland" category, due to small number of participants residing in Northern Ireland. Scotland was chosen as the geographically closest other UK region.

Equivalent data for education level and residential address immediately prior to the COVID-19 pandemic was not available and responses were assumed to represent pre-pandemic statuses.

Information on pre-pandemic employment status was self-reported in the May, 2021 CSSB questionnaire in the question "Which one of these best describes what you were doing before the COVID-19 pandemic? If you were doing more than one activity, please choose the activity you spent the most time doing". Due to small numbers of individuals, "Other" was a heterogeneous group comprising "In unpaid/voluntary work", "In education at school/college/university, or in an apprenticeship", "Looking after home or family", and "Other" original response options.

Current employment status was self-reported in the August, 2022 CSSB questionnaire, with "Other" options grouped as for the pre-pandemic equivalent.

Current yearly gross household income was self-reported in the August, 2022 CSSB questionnaire with the question "What is the total yearly income before tax received by your household? This includes all those who can earn and live in the same household as yourself."

*Health characteristics*

Pre-pandemic general health was self-reported in the May, 2021 CSSB questionnaire with the question "In general, in the 3 months before the COVID-19 outbreak in March 2020, would you say your health was...".

Body mass index (BMI) was derived from self-reported height and weight collected at CSSB consent.

Frailty, a measure of age-related decline in physiological reserve and function [6], was measured using the PRISMA-7 scale [7], collected at registration with the CSS app.

Number of physical health conditions was measured from 6 self-reported conditions (asthma, cancer, diabetes, heart disease, lung disease, kidney disease) collected at registration for the CSS app.

Number of mental health conditions was measured from self-reported diagnoses of 16 conditions (Generalised anxiety order; Panic disorder; Specific phobias; Obsessive compulsive disorder; Post-traumatic stress disorder; Social anxiety disorder; Agoraphobia; Depression; Attention deficit or attention deficit and hyperactivity disorder; Autism, Asperger's or autistic spectrum disorder; Eating disorder (e.g. bulimia nervosa; anorexia nervosa; psychological over-eating or binge-eating), Personality disorder; Mania, hypomania, bipolar or manic depression; Schizophrenia; Substance use disorder; Any other type of psychosis or psychotic illness) collected in a February, 2021 CSS questionnaire.

Equivalent data used to derive BMI, frailty and number of physical health conditions representing status immediately prior to the COVID-19 pandemic was not available and responses were assumed to represent pre-pandemic statuses.

*COVID-19 illness characteristics*

COVID-19 illness characteristics were derived from self-report in the August, 2022 CSSB questionnaire. COVID-19 infection history was measured with the question, "How many times do you think you have ever had COVID-19? Please include now if you think you currently have COVID-19 symptoms but have not confirmed it.". For each reported infection, participants were asked what evidence supported their infection ("How do/did you know you had it (COVID-19)?"), the start date of the infection/illness (“When do you think you had COVID-19? Please use your best estimate if you can't remember the exact date.”), the duration of COVID-19 symptoms (“How long did you have or have you had continuous symptoms?”). Where multiple infections were reported, information was obtained for the single infection with the longest reported symptom duration, then earliest date of infection start, then strongest evidence of infection.

Infection period was derived from self-reported date of infection start. Periods were defined based on changes in dominant SARS-CoV-2 variant from COG-UK Mutation Explorer data available at https://sars2.cvr.gla.ac.uk/cog-uk/.

Severity of acute COVID-19 illness was assessed based on whether an individual accessed urgent care, derived from the question, “What type of medical help did you access WITHIN the first 4 weeks of (/MORE THAN 4 weeks after) the start of your symptoms that you think may have been caused by COVID-19?”. Selection of options “Visited A&E or walk-in centre” or “Called an ambulance” were taken as indicating use of urgent care.

Long COVID diagnosis status was measured by the question, “Have you ever received a diagnosis of long COVID or post-COVID syndrome?”, with the following response options: “Yes”, “No, but I do believe I have or have had Long COVID”, and “No, and I do not believe I have or have had Long COVID”.

Self-perceived recovery following COVID-19 was measured by the question, “Thinking about the last or only episode of COVID-19 you have had, have you now recovered and are back to normal?”, with the following response options: “Yes, I am back to normal”, “No, I still have some or all my symptoms”.

#### Sample selection flow diagram

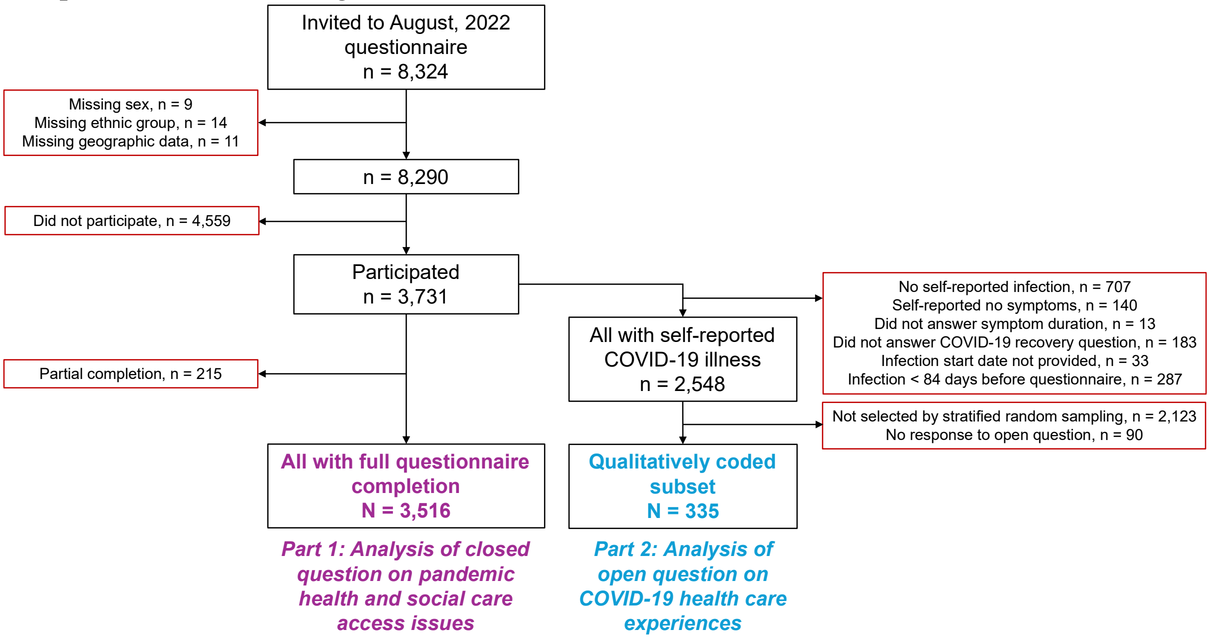

*Figure S 1. Sample selection flow diagram. Exclusions are identified in red boxes.*

Full directed acyclic graph & daggity code

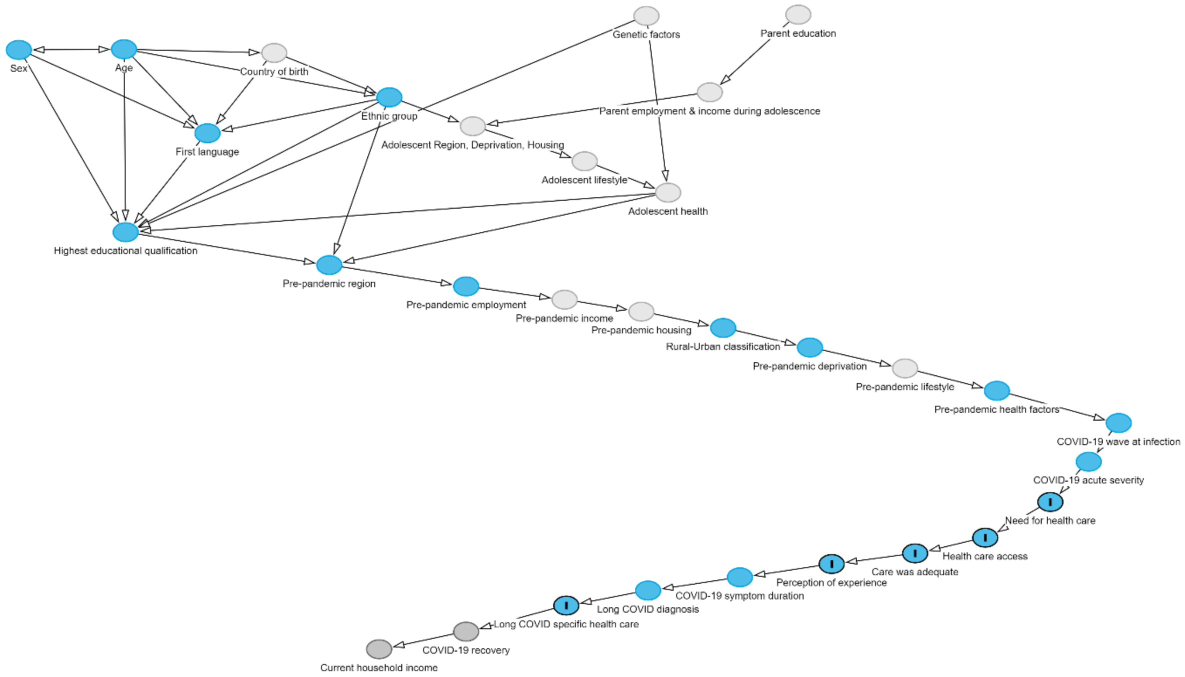

*Figure S 2. Proposed directed acyclic graph (DAG) used to generate adjustment variable sets for models estimating the total causal effect of exposure variables on health care access and experiences. The proposed DAG is 'saturated', in that each variable is hypothesised to be caused by all earlier variables, but arcs to all subsequent variables are not all drawn for clarity. Outcomes in analyses are marked with 'I', and unobserved potential confounding variables are coloured in light grey.*

Below is code to recreate the full DAG using daggity software:

```
dag {

"Adolescent Region, Deprivation, Housing" [latent,pos="-0.855,-1.118"]

"Adolescent health" [latent,pos="-0.738,-0.813"]

"Adolescent lifestyle" [latent,pos="-0.788,-0.957"]

"COVID-19 acute severity" [pos="-0.486,0.424"]

"COVID-19 recovery" [pos="-0.859,1.205"]

"COVID-19 symptom duration" [pos="-0.695,0.955"]

"COVID-19 wave at infection" [pos="-0.468,0.246"]

"Care was adequate" [outcome,pos="-0.590,0.845"]

"Country of birth" [latent,pos="-0.974,-1.455"]

"Current household income" [pos="-0.911,1.286"]

"Ethnic group" [pos="-0.905,-1.251"]

"First language" [pos="-1.014,-1.086"]

"Genetic factors" [latent,pos="-0.751,-1.625"]
```

"Health care access" [outcome,pos="-0.548,0.773"]

"Highest educational qualification" [pos="-1.063,-0.631"]

"Long COVID diagnosis" [pos="-0.750,1.016"]

"Long COVID specific health care" [outcome,pos="-0.799,1.085"]

"Need for health care" [outcome,pos="-0.509,0.609"]

"Parent education" [latent,pos="-0.660,-1.631"]

"Parent employment & income during adolescence" [latent,pos="-0.713,-1.274"]

"Perception of experience" [outcome,pos="-0.640,0.894"]

"Pre-pandemic deprivation" [pos="-0.653,-0.101"]

"Pre-pandemic employment" [pos="-0.859,-0.382"]

"Pre-pandemic health factors" [pos="-0.541,0.098"]

"Pre-pandemic housing" [latent,pos="-0.754,-0.266"]

"Pre-pandemic income" [latent,pos="-0.800,-0.321"]

"Pre-pandemic lifestyle" [latent,pos="-0.596,-0.005"]

"Pre-pandemic region" [pos="-0.941,-0.480"]

"Rural-Urban classification" [pos="-0.705,-0.191"]

"border bottom left" [pos="-1.145,1.601"]

"border bottom right" [pos="-0.418,1.577"]

"border top left" [pos="-1.157,-1.865"]

"border top right" [pos="-0.415,-1.865"]

Age [pos="-1.064,-1.472"]

Sex [pos="-1.127,-1.469"]

"Adolescent Region, Deprivation, Housing" -> "Adolescent lifestyle"

"Adolescent health" -> "Highest educational qualification"

"Adolescent health" -> "Pre-pandemic region"

"Adolescent lifestyle" -> "Adolescent health"

"COVID-19 acute severity" -> "Need for health care"

"COVID-19 recovery" -> "Current household income"

"COVID-19 symptom duration" -> "Long COVID diagnosis"

"COVID-19 wave at infection" -> "COVID-19 acute severity"

"Care was adequate" -> "Perception of experience"

"Country of birth" -> "Ethnic group"

```
"Country of birth" -> "First language"
"Ethnic group" -> "Adolescent Region, Deprivation, Housing"
"Ethnic group" -> "First language"
"Ethnic group" -> "Highest educational qualification"
"Ethnic group" -> "Pre-pandemic region"
"First language" -> "Highest educational qualification"
"Genetic factors" -> "Adolescent health"
"Genetic factors" -> "Highest educational qualification"
"Health care access" -> "Care was adequate"
"Highest educational qualification" -> "Pre-pandemic region"
"Long COVID diagnosis" -> "Long COVID specific health care"
"Long COVID specific health care" -> "COVID-19 recovery"
"Need for health care" -> "Health care access"
"Parent education" -> "Parent employment & income during adolescence"
"Parent employment & income during adolescence" -> "Adolescent Region, Deprivation, Housing"
"Perception of experience" -> "COVID-19 symptom duration"
"Pre-pandemic deprivation" -> "Pre-pandemic lifestyle"
"Pre-pandemic employment" -> "Pre-pandemic income"
"Pre-pandemic health factors" -> "COVID-19 wave at infection"
"Pre-pandemic housing" -> "Rural-Urban classification"
"Pre-pandemic income" -> "Pre-pandemic housing"
"Pre-pandemic lifestyle" -> "Pre-pandemic health factors"
"Pre-pandemic region" -> "Pre-pandemic employment"
"Rural-Urban classification" -> "Pre-pandemic deprivation"
Age -> "Country of birth"
Age -> "Ethnic group"
Age -> "First language"
Age -> "Highest educational qualification"
Age <-> Sex
Sex -> "First language"
Sex -> "Highest educational qualification"
}
```

#### Inverse probability weight models summary

*Table S 1. Summary of models used to derive inverse probability weights used in models estimating associations with health and social care access issues and COVID-19 health care experiences.*

| Model | Sample model trained on | Model outcome | Variable set | AUC-ROC score |
|---|---|---|---|---|
| 1 | All invited to August 2022 questionnaire | Full completion of August 2022 questionnaire | Age group, ethnic group, CSSB recruitment group, number of mental health conditions, local area deprivation, PRISMA-7 frailty score, number of non-responses to previous CSSB studies | 0.84 |
| 2 | All August 2022 questionnaire participants | All with self-reported COVID-19 illness | Sex, first language, education level, pre-pandemic employment status, pre-pandemic general health, worked as health care professional, UK region, PRISMA-7 frailty score | 0.65 |
| 3 | Qualitatively coded subset (including those with no response to open question on COVID-19 health care experiences) | Non-zero response to open question on COVID-19 health care experiences | Age group, sex, ethnic group, pre-pandemic general health, BMI | 0.76 |
| 4 | Qualitatively coded subset (excluding non-zero responses to open question) | Care needed for COVID-19 | Sex, local area deprivation, pre-pandemic general health, BMI | 0.71 |

Inverse probability weights used to estimate associations with health and social care access issues (analysis part 1) were derived from model 1 only.

Inverse probability weights used to estimate associations with need for health care for COVID-19 (analysis part 2) were generated from multiplication of IPWs derived from model 1, model 2, and model 3, i.e. IPW(model 1) x IPW(model 2) x IPW(model 3).

Inverse probability weights used to estimate associations with COVID-19 health care ease of access, perception of experiences, and access to long COVID clinic or specialist care (analysis part 2), were generated from multiplication of IPWs derived from model 1-4, i.e. IPW(model 1) x IPW(model 2) x IPW(model 3) x IPW(model 4).

#### Extended sample characteristics and multivariable regression model results testing association with individual factors

*Table S 2. Extended sample characteristics and results of multivariable poisson and linear regression models. Relative risk ratios (poisson models) and coefficients (linear models), 95% confidence intervals (CI) and p-values adjusted for multiple testing (Benjamini/Hochberg false discovery rate correction) are presented for multivariable regression models estimating associations between health & social care access issues and exposures of interest, after adjustment as appropriate from the hypothesised directed acyclic graph (DAG), and weighting for inverse probability of questionnaire completion. Adjusted p-values < 0.05 are highlighted in bold. Figures for group sizes of less than 5 are suppressed.*

| | | Group size | | One or more health & social care access issues | | Poisson regression, individual factors | | | | Linear regression, individual factors | | | |
|---|---|---|---|---|---|---|---|---|---|---|---|---|---|
| Domain | Variable | N | % | N | % | Relative risk ratio | 95% CI (lower) | 95% CI (upper) | P-value (adjusted) | Coefficient | 95% CI (lower) | 95% CI (upper) | P-value (adjusted) |
| | **Total** | 3516 | | 999 | 28.4% | | | | | | | | |
| **Health & social care access issues during the COVID-19 pandemic** | **Health & social care access issue type** | | | | | | | | | | | | |
| | Unable to access required medication | 144 | 4.1% | | | | | | | | | | |
| | Unable to access health services in the community | 827 | 23.5% | | | | | | | | | | |
| | Unable to access the community social care services or voluntary sector support needed | 73 | 2.1% | | | | | | | | | | |
| | Unable to access inpatient or outpatient appointments booked at a hospital | 388 | 11.0% | | | | | | | | | | |
| | Unable to access appointment for cognitive behaviour therapy, counselling, or psychological therapy | 127 | 3.6% | | | | | | | | | | |
| | **Health & social care access issue count** | | | | | | | | | | | | |
| | None | 2517 | 71.6% | | | | | | | | | | |
| | One | 624 | 17.7% | | | | | | | | | | |
| | Two | 251 | 7.1% | | | | | | | | | | |
| | Three | 81 | 2.3% | | | | | | | | | | |
| | Four | 25 | 0.7% | | | | | | | | | | |
| | Five | 18 | 0.5% | | | | | | | | | | |
| **Individual pre-pandemic demographics** | **Age (median, interquartile range)** | 59 (52-65) | | 58 (52-65) | | | | | | | | | |
| | **Age group (years)** | | | | | | | | | | | | |
| | 18-39 | 186 | 5.3% | 47 | 25.3% | 0.84 | 0.61 | 1.14 | 0.438 | 0.01 | -0.11 | 0.13 | 1.000 |
| | 40-49 | 493 | 14.0% | 148 | 30.0% | 0.94 | 0.78 | 1.14 | 0.705 | -0.07 | -0.16 | 0.01 | 0.226 |
| | 50-59 (reference) | 1144 | 32.5% | 362 | 31.6% | 1.00 | | | | 0.00 | | | |
| | 60-69 | 1254 | 35.7% | 322 | 25.7% | 0.86 | 0.73 | 1.00 | 0.151 | -0.13 | -0.20 | -0.06 | **0.003** |
| | ≥ 70 | 439 | 12.5% | 120 | 27.3% | 0.92 | 0.74 | 1.13 | 0.600 | -0.07 | -0.18 | 0.04 | 0.371 |
| | **Sex** | | | | | | | | | | | | |
| | Female (reference) | 2840 | 80.8% | 843 | 29.7% | 1.00 | | | | 0.00 | | | |
| | Male | 676 | 19.2% | 156 | 23.1% | 0.85 | 0.71 | 1.02 | 0.223 | -0.11 | -0.18 | -0.03 | **0.030** |
| | **Ethnic group** | | | | | | | | | | | | |
| | Asian/Asian British | 18 | 0.5% | <5 | - | 0.91 | 0.37 | 2.24 | 0.948 | -0.02 | -0.34 | 0.29 | 1.000 |
| | Black/Black British | 15 | 0.4% | <5 | - | 0.48 | 0.15 | 1.51 | 0.386 | -0.29 | -0.75 | 0.18 | 0.407 |

| | | | | | | | | | | | | | |
|---|---|---|---|---|---|---|---|---|---|---|---|---|---|
| | Mixed/Multiple | 29 | 0.8% | 9 | 31.0% | 0.86 | 0.44 | 1.69 | 0.808 | -0.12 | -0.41 | 0.17 | 0.640 |
| | Other | 38 | 1.1% | 11 | 28.9% | 0.96 | 0.53 | 1.73 | 0.985 | -0.10 | -0.36 | 0.15 | 0.640 |
| | White (reference) | 3416 | 97.2% | 972 | 28.5% | 1.00 | | | | 0.00 | | | |
| | **First language** | | | | | | | | | | | | |
| | English (reference) | 3437 | 97.8% | 976 | 28.4% | 1.00 | | | | 0.00 | | | |
| | Other | 76 | 2.2% | 23 | 30.3% | 0.98 | 0.63 | 1.52 | 1.000 | 0.03 | -0.17 | 0.24 | 0.996 |
| | Prefer not to answer/not stated | <5 | - | 0 | 0.0% | | | | | | | | |
| | **Highest educational qualification** | | | | | | | | | | | | |
| | Prefer not to answer/not stated | 40 | 1.1% | 13 | 32.5% | 1.51 | 0.91 | 2.52 | 0.273 | 0.41 | 0.15 | 0.66 | **0.008** |
| | Did not complete secondary school | 25 | 0.7% | <5 | - | 0.78 | 0.29 | 2.10 | 0.761 | 0.02 | -0.32 | 0.37 | 1.000 |
| | GCSE or GNVQ or equivalent | 412 | 11.7% | 139 | 33.7% | 1.23 | 1.01 | 1.51 | 0.135 | 0.05 | -0.04 | 0.15 | 0.458 |
| | A-Levels or advanced GNVQ or equivalent | 587 | 16.7% | 183 | 31.2% | 1.14 | 0.95 | 1.37 | 0.317 | 0.01 | -0.08 | 0.10 | 1.000 |
| | University degree (reference) | 1269 | 36.1% | 341 | 26.9% | 1.00 | | | | 0.00 | | | |
| | Postgraduate degree or higher | 1014 | 28.8% | 272 | 26.8% | 0.91 | 0.77 | 1.08 | 0.458 | -0.06 | -0.13 | 0.02 | 0.256 |
| | PhD | 169 | 4.8% | 47 | 27.8% | 1.06 | 0.77 | 1.46 | 0.831 | -0.04 | -0.19 | 0.11 | 0.780 |
| | **UK Region** | | | | | | | | | | | | |
| | East Midlands | 227 | 6.5% | 74 | 32.6% | 1.24 | 0.93 | 1.65 | 0.301 | 0.13 | -0.01 | 0.26 | 0.157 |
| | East of England | 377 | 10.7% | 113 | 30.0% | 1.23 | 0.95 | 1.57 | 0.273 | 0.10 | -0.01 | 0.22 | 0.183 |
| | London (reference) | 593 | 16.9% | 163 | 27.5% | 1.00 | | | | 0.00 | | | |
| | North East | 109 | 3.1% | 29 | 26.6% | 1.11 | 0.76 | 1.64 | 0.733 | 0.01 | -0.18 | 0.21 | 1.000 |
| | North West | 342 | 9.7% | 95 | 27.8% | 0.96 | 0.73 | 1.25 | 0.871 | -0.07 | -0.19 | 0.05 | 0.443 |
| | Scotland & Northern Ireland | 178 | 5.1% | 54 | 30.3% | 1.17 | 0.85 | 1.61 | 0.501 | 0.02 | -0.13 | 0.18 | 1.000 |
| | South East | 693 | 19.7% | 195 | 28.1% | 1.11 | 0.89 | 1.38 | 0.547 | 0.05 | -0.04 | 0.15 | 0.468 |
| | South West | 360 | 10.2% | 90 | 25.0% | 1.05 | 0.81 | 1.38 | 0.829 | 0.00 | -0.12 | 0.12 | 1.000 |
| | Wales | 180 | 5.1% | 53 | 29.4% | 0.89 | 0.63 | 1.26 | 0.687 | -0.11 | -0.26 | 0.04 | 0.313 |
| | West Midlands | 223 | 6.3% | 67 | 30.0% | 1.27 | 0.95 | 1.70 | 0.273 | 0.05 | -0.08 | 0.19 | 0.640 |
| | Yorkshire and The Humber | 234 | 6.7% | 66 | 28.2% | 1.14 | 0.85 | 1.53 | 0.562 | -0.07 | -0.20 | 0.07 | 0.538 |
| | **Pre-pandemic employment status** | | | | | | | | | | | | |
| | Employed (reference) | 1777 | 50.5% | 530 | 29.8% | 1.00 | | | | 0.00 | | | |
| | Self-employed | 390 | 11.1% | 106 | 27.2% | 1.06 | 0.86 | 1.31 | 0.740 | 0.06 | -0.04 | 0.15 | 0.429 |
| | Unemployed | 15 | 0.4% | 6 | 40.0% | 1.53 | 0.87 | 2.69 | 0.301 | 0.45 | -0.07 | 0.97 | 0.203 |
| | Permanently or long-term sick or disabled | 46 | 1.3% | 35 | 76.1% | 2.70 | 2.27 | 3.21 | **7.32E-27** | 1.90 | 1.63 | 2.16 | **0.000** |
| | Retired | 928 | 26.4% | 206 | 22.2% | 0.75 | 0.61 | 0.93 | **0.043** | -0.10 | -0.19 | 0.00 | 0.136 |
| | Other | 287 | 8.2% | 87 | 30.3% | 1.00 | 0.80 | 1.25 | 1.000 | 0.05 | -0.07 | 0.17 | 0.640 |
| | Unknown | 73 | 2.1% | 29 | 39.7% | 1.43 | 1.03 | 1.97 | 0.117 | 0.45 | 0.30 | 0.60 | **1.06E-07** |
| | **Rural-Urban classification** | | | | | | | | | | | | |
| | Rural | 864 | 24.6% | 227 | 26.3% | 0.87 | 0.74 | 1.02 | 0.223 | -0.08 | -0.15 | -0.01 | 0.107 |
| | Urban (reference) | 2652 | 75.4% | 772 | 29.1% | 1.00 | | | | 0.00 | | | |
| | **Local area deprivation** | | | | | | | | | | | | |
| | IMD Decile 1 (most deprived 10%) | 82 | 2.3% | 37 | 45.1% | 1.73 | 1.21 | 2.47 | **0.020** | 0.24 | 0.04 | 0.43 | 0.060 |
| | IMD Decile 2 | 129 | 3.7% | 42 | 32.6% | 1.32 | 0.95 | 1.85 | 0.270 | 0.13 | -0.03 | 0.30 | 0.250 |
| | IMD Decile 3 | 185 | 5.3% | 57 | 30.8% | 1.27 | 0.93 | 1.75 | 0.301 | 0.12 | -0.02 | 0.26 | 0.226 |
| | IMD Decile 4 | 255 | 7.3% | 90 | 35.3% | 1.57 | 1.21 | 2.05 | **0.007** | 0.24 | 0.12 | 0.37 | **0.001** |
| | IMD Decile 5 | 314 | 8.9% | 95 | 30.3% | 1.30 | 0.99 | 1.71 | 0.183 | 0.12 | 0.01 | 0.24 | 0.113 |
| | IMD Decile 6 | 406 | 11.5% | 122 | 30.0% | 1.32 | 1.03 | 1.69 | 0.117 | 0.11 | 0.00 | 0.22 | 0.138 |
| | IMD Decile 7 | 428 | 12.2% | 129 | 30.1% | 1.59 | 1.25 | 2.01 | **0.002** | 0.29 | 0.18 | 0.40 | **1.59E-06** |

| | | | | | | | | | | | | | |
|---|---|---|---|---|---|---|---|---|---|---|---|---|---|
| | IMD Decile 8 | 495 | 14.1% | 131 | 26.5% | 1.17 | 0.92 | 1.50 | 0.377 | 0.10 | -0.01 | 0.20 | 0.159 |
| | IMD Decile 9 | 563 | 16.0% | 150 | 26.6% | 1.16 | 0.91 | 1.46 | 0.386 | 0.08 | -0.02 | 0.18 | 0.256 |
| | IMD Decile 10 (least deprived 10%) (reference) | 659 | 18.7% | 146 | 22.2% | 1.00 | | | | 0.00 | | | |
| **Socio-demographics during pandemic** | **Current household income** | | | | | | | | | | | | |
| | Prefer not to answer/not stated | 467 | 13.3% | 146 | 31.3% | 1.15 | 0.92 | 1.43 | 0.386 | 0.09 | -0.02 | 0.19 | 0.226 |
| | Less than £20,000 | 318 | 9.0% | 127 | 39.9% | 1.19 | 0.95 | 1.49 | 0.284 | 0.20 | 0.07 | 0.32 | **0.008** |
| | £20,000-£29,999 | 348 | 9.9% | 103 | 29.6% | 1.08 | 0.85 | 1.38 | 0.687 | 0.05 | -0.07 | 0.17 | 0.640 |
| | £30,000-£39,999 | 425 | 12.1% | 128 | 30.1% | 1.15 | 0.92 | 1.43 | 0.386 | 0.13 | 0.02 | 0.24 | 0.059 |
| | £40,000-£49,999 | 415 | 11.8% | 111 | 26.7% | 0.87 | 0.68 | 1.11 | 0.428 | 0.00 | -0.10 | 0.11 | 1.000 |
| | £50,000-£74,999 (reference) | 649 | 18.5% | 178 | 27.4% | 1.00 | | | | 0.00 | | | |
| | £75,000-£99,999 | 378 | 10.8% | 106 | 28.0% | 1.12 | 0.88 | 1.43 | 0.526 | 0.03 | -0.08 | 0.14 | 0.780 |
| | £100,000 or more | 516 | 14.7% | 100 | 19.4% | 0.71 | 0.54 | 0.92 | **0.043** | -0.12 | -0.21 | -0.02 | 0.064 |
| **Pre-pandemic health factors** | **Pre-pandemic health** | | | | | | | | | | | | |
| | Poor | 94 | 2.7% | 52 | 55.3% | 1.73 | 1.30 | 2.30 | **0.002** | 0.44 | 0.24 | 0.65 | **1.72E-04** |
| | Fair | 286 | 8.1% | 118 | 41.3% | 1.64 | 1.35 | 2.00 | **2.20E-05** | 0.38 | 0.26 | 0.50 | **4.27E-08** |
| | Good | 768 | 21.8% | 271 | 35.3% | 1.46 | 1.24 | 1.71 | **7.27E-05** | 0.19 | 0.10 | 0.27 | **1.17E-04** |
| | Very good (reference) | 1265 | 36.0% | 299 | 23.6% | 1.00 | | | | 0.00 | | | |
| | Excellent | 663 | 18.9% | 129 | 19.5% | 0.84 | 0.68 | 1.05 | 0.284 | 0.01 | -0.08 | 0.09 | 1.000 |
| | Unknown | 440 | 12.5% | 130 | 29.5% | 1.15 | 0.93 | 1.42 | 0.377 | 0.13 | 0.05 | 0.21 | **0.008** |
| | **Body mass index** | | | | | | | | | | | | |
| | < 18.5 kg/m^2 | 42 | 1.2% | 12 | 28.6% | 1.39 | 0.82 | 2.37 | 0.386 | 0.34 | 0.07 | 0.61 | 0.054 |
| | 18.5-25 kg/m^2 (reference) | 1536 | 43.7% | 364 | 23.7% | 1.00 | | | | 0.00 | | | |
| | 25-30 kg/m^2 | 1106 | 31.5% | 320 | 28.9% | 1.21 | 1.04 | 1.42 | 0.076 | 0.14 | 0.07 | 0.21 | **3.88E-04** |
| | ≥ 30 kg/m^2 | 826 | 23.5% | 300 | 36.3% | 1.42 | 1.21 | 1.66 | **1.83E-04** | 0.18 | 0.10 | 0.25 | **6.84E-05** |
| | Unknown | 6 | 0.2% | <5 | - | | | | | | | | |
| | **PRISMA-7 Frailty score** | | | | | | | | | | | | |
| | 0-2, below threshold (reference) | 3333 | 94.8% | 910 | 27.3% | 1.00 | | | | 0.00 | | | |
| | 3-7, above threshold | 183 | 5.2% | 89 | 48.6% | 1.70 | 1.39 | 2.07 | **7.30E-06** | 0.44 | 0.30 | 0.59 | **4.27E-08** |
| | **Number of physical health conditions** | | | | | | | | | | | | |
| | None (reference) | 2700 | 76.8% | 712 | 26.4% | 1.00 | | | | 0.00 | | | |
| | One | 705 | 20.1% | 236 | 33.5% | 1.18 | 1.02 | 1.36 | 0.116 | 0.11 | 0.03 | 0.18 | **0.017** |
| | Two or more | 111 | 3.2% | 51 | 45.9% | 1.73 | 1.35 | 2.20 | **1.83E-04** | 0.55 | 0.39 | 0.72 | **7.22E-09** |
| | **Number of mental health conditions (February 2021)** | | | | | | | | | | | | |
| | None (reference) | 2190 | 62.3% | 544 | 24.8% | 1.00 | | | | 0.00 | | | |
| | One | 516 | 14.7% | 178 | 34.5% | 1.15 | 0.97 | 1.36 | 0.273 | 0.05 | -0.03 | 0.14 | 0.413 |
| | Two | 163 | 4.6% | 70 | 42.9% | 1.39 | 1.11 | 1.74 | **0.030** | 0.18 | 0.04 | 0.32 | 0.054 |
| | Three or more | 78 | 2.2% | 40 | 51.3% | 1.49 | 1.10 | 2.00 | **0.043** | 0.50 | 0.30 | 0.70 | **1.73E-05** |
| | Unknown | 569 | 16.2% | 167 | 29.3% | 1.10 | 0.92 | 1.31 | 0.472 | 0.09 | 0.02 | 0.16 | **0.049** |
| **COVID-19 illness characteristics** | **COVID-19 history** | | | | | | | | | | | | |
| | No history of COVID-19 | 598 | 17.0% | 182 | 30.4% | | | | | | | | |
| | COVID-19 with positive antigen test | 2078 | 59.1% | 576 | 27.7% | | | | | | | | |
| | COVID-19 with positive antibody test | 504 | 14.3% | 133 | 26.4% | | | | | | | | |
| | COVID-19 based on medical advice | 78 | 2.2% | 37 | 47.4% | | | | | | | | |

| | | | | | |
|---|---|---|---|---|---|
| | COVID-19 based on strong personal suspicion | 245 | 7.0% | 68 | 27.8% |
| | Unsure had COVID-19 | 13 | 0.4% | <5 | - |
| | **Infection period** | | | | |
| | N/A - no COVID-19 | 598 | 17.0% | 182 | 30.4% |
| | COVID-19 start date unknown | 29 | 0.8% | 7 | 24.1% |
| | Before 2020-12-08 (pre-vaccination, wild-type dominant) (reference) | 1482 | 42.2% | 434 | 29.3% |
| | 2020-12-08 to 2021-04-25 (alpha-variant dominant) | 271 | 7.7% | 97 | 35.8% |
| | 2021-04-25 to 2021-12-08 (delta-variant dominant) | 146 | 4.2% | 31 | 21.2% |
| | After 2021-12-08 (omicron-variant dominant) | 990 | 28.2% | 248 | 25.1% |
| | **Urgent care accessed during COVID-19 illness** | | | | |
| | N/A - no COVID-19 | 598 | 17.0% | 182 | 30.4% |
| | No (reference) | 2547 | 72.4% | 685 | 26.9% |
| | Yes | 222 | 6.3% | 101 | 45.5% |
| | Unknown | 149 | 4.2% | 31 | 20.8% |
| | **Symptom duration (retrospective self-report)** | | | | |
| | N/A - no COVID-19 | 598 | 17.0% | 182 | 30.4% |
| | No symptoms | 140 | 4.0% | 27 | 19.3% |
| | Less than 2 weeks (reference) | 1110 | 31.6% | 240 | 21.6% |
| | 2 to 4 weeks | 635 | 18.1% | 179 | 28.2% |
| | 4 to 12 weeks | 360 | 10.2% | 130 | 36.1% |
| | 3 to 6 months | 150 | 4.3% | 57 | 38.0% |
| | 6 to 12 months | 118 | 3.4% | 35 | 29.7% |
| | 12 to 18 months | 76 | 2.2% | 22 | 28.9% |
| | 18 to 24 months | 136 | 3.9% | 64 | 47.1% |
| | 24 months or more | 184 | 5.2% | 59 | 32.1% |
| | Unknown | 9 | 0.3% | <5 | - |
| | **Long COVID diagnosis status** | | | | |
| | N/A - no COVID-19 | 598 | 17.0% | 182 | 30.4% |
| | No diagnosis, and does not believe has/had long COVID | 1647 | 46.8% | 388 | 23.6% |
| | No diagnosis, but believe have/had long COVID (reference) | 789 | 22.4% | 267 | 33.8% |
| | Diagnosed with Long COVID | 332 | 9.4% | 130 | 39.2% |
| | Unknown | 150 | 4.3% | 32 | 21.3% |
| | **Self-perceived COVID-19 recovery** | | | | |
| | N/A - no COVID-19 | 598 | 17.0% | 182 | 30.4% |
| | Recovered | 1884 | 53.6% | 458 | 24.3% |
| | Not recovered | 873 | 24.8% | 325 | 37.2% |
| | Unknown | 161 | 4.6% | 34 | 21.1% |

#### MAIHDA intersectional mixed-effects model results tables (corresponding to main text Figure 3)

##### *Stratum-level predicted probabilities*

*Table S 3. Stratum-level predictions of health & social care access issues from MAIHDA models (corresponding to main text Figure 3). Unadjusted mean number of issues and proportion of one or more issues are given alongside model predictions (with approximate 95% confidence intervals) from MAIHDA mixed-effects regression models for combinations of sex, education level and local area deprivation. Confidence intervals are approximate only because the model assumes no sampling covariability between the regression coefficients and stratum random effects. Models included participant weighting for inverse probability of questionnaire participation. Strata labels: F = Female, M = Male; NoDegree = Less than degree level education (including not stated/prefer not to say), Undergrad = Undergraduate degree level, Postgrad = Postgraduate degree level or higher; IMD1-IMD5 = Index of Multiple Deprivation Quintile 1 to 5, where 1 is most deprived 20% of areas, and 5 is least deprived 20%.*

| Stratum | Stratum size (N) | Unadjusted mean number of health & social care issues | Unadjusted proportion with one or more issues (%) | Predicted mean number of issues | Lower 95% CI | Upper 95% CI | Predicted probability of one or more issues (%) | Lower 95% CI | Upper 95% CI |
|---|---|---|---|---|---|---|---|---|---|
| F + NoDegree + IMD1 | 77 | 0.69 | 42.9 | 0.66 | 0.45 | 0.87 | 42.9 | 31.9 | 54.6 |
| F + NoDegree + IMD2 | 113 | 0.54 | 31.0 | 0.57 | 0.38 | 0.77 | 36.3 | 26.7 | 47.3 |
| F + NoDegree + IMD3 | 167 | 0.42 | 29.9 | 0.44 | 0.25 | 0.62 | 30.2 | 22.6 | 40.2 |
| F + NoDegree + IMD4 | 233 | 0.50 | 32.6 | 0.55 | 0.37 | 0.71 | 34.6 | 26.2 | 44.1 |
| F + NoDegree + IMD5 | 268 | 0.45 | 30.2 | 0.47 | 0.31 | 0.64 | 29.9 | 22.2 | 38.4 |
| F + Undergrad + IMD1 | 48 | 0.58 | 33.3 | 0.55 | 0.30 | 0.78 | 33.4 | 22.5 | 47.1 |
| F + Undergrad + IMD2 | 131 | 0.66 | 38.9 | 0.61 | 0.41 | 0.79 | 35.2 | 26.3 | 45.6 |
| F + Undergrad + IMD3 | 215 | 0.55 | 34.0 | 0.53 | 0.37 | 0.70 | 32.2 | 23.8 | 41.8 |
| F + Undergrad + IMD4 | 278 | 0.52 | 30.6 | 0.57 | 0.40 | 0.73 | 31.2 | 23.5 | 39.9 |
| F + Undergrad + IMD5 | 356 | 0.31 | 19.4 | 0.34 | 0.18 | 0.49 | 21.1 | 15.6 | 28.0 |
| F + Postgrad + IMD1 | 50 | 0.54 | 34.0 | 0.49 | 0.25 | 0.72 | 31.1 | 21.5 | 43.5 |
| F + Postgrad + IMD2 | 126 | 0.45 | 28.6 | 0.43 | 0.24 | 0.63 | 27.1 | 19.3 | 36.9 |
| F + Postgrad + IMD3 | 213 | 0.56 | 31.0 | 0.48 | 0.31 | 0.66 | 27.7 | 20.2 | 37.4 |
| F + Postgrad + IMD4 | 235 | 0.46 | 29.4 | 0.51 | 0.34 | 0.67 | 29.1 | 21.5 | 37.3 |
| F + Postgrad + IMD5 | 330 | 0.37 | 26.1 | 0.37 | 0.21 | 0.53 | 24.0 | 17.8 | 31.2 |
| M + NoDegree + IMD1 | 18 | 0.78 | 50.0 | 0.56 | 0.32 | 0.81 | 37.8 | 26.6 | 51.6 |
| M + NoDegree + IMD2 | 24 | 0.71 | 50.0 | 0.55 | 0.31 | 0.78 | 36.5 | 25.5 | 49.9 |
| M + NoDegree + IMD3 | 42 | 0.40 | 28.6 | 0.46 | 0.25 | 0.67 | 28.9 | 20.0 | 39.6 |
| M + NoDegree + IMD4 | 49 | 0.20 | 16.3 | 0.36 | 0.14 | 0.56 | 25.6 | 17.6 | 35.7 |
| M + NoDegree + IMD5 | 73 | 0.45 | 31.5 | 0.42 | 0.21 | 0.63 | 28.2 | 19.7 | 38.8 |
| M + Undergrad + IMD1 | 9 | 0.33 | 33.3 | 0.46 | 0.22 | 0.72 | 31.0 | 20.1 | 44.2 |
| M + Undergrad + IMD2 | 25 | 0.32 | 24.0 | 0.44 | 0.19 | 0.66 | 28.5 | 19.2 | 40.2 |
| M + Undergrad + IMD3 | 44 | 0.34 | 22.7 | 0.43 | 0.21 | 0.65 | 25.8 | 17.8 | 36.5 |
| M + Undergrad + IMD4 | 68 | 0.31 | 17.6 | 0.42 | 0.22 | 0.62 | 23.8 | 16.6 | 32.7 |
| M + Undergrad + IMD5 | 95 | 0.21 | 16.8 | 0.28 | 0.09 | 0.47 | 20.2 | 13.9 | 28.1 |
| M + Postgrad + IMD1 | 9 | 0.22 | 11.1 | 0.46 | 0.21 | 0.71 | 28.6 | 18.7 | 41.7 |
| M + Postgrad + IMD2 | 21 | 0.48 | 33.3 | 0.40 | 0.14 | 0.67 | 28.0 | 17.9 | 41.0 |
| M + Postgrad + IMD3 | 39 | 0.21 | 15.4 | 0.27 | 0.07 | 0.50 | 20.6 | 13.8 | 29.6 |
| M + Postgrad + IMD4 | 60 | 0.17 | 16.7 | 0.27 | 0.07 | 0.48 | 21.1 | 14.3 | 29.5 |
| M + Postgrad + IMD5 | 100 | 0.31 | 21.0 | 0.25 | 0.06 | 0.44 | 18.7 | 12.8 | 26.7 |

#### *MAIHDA model parameter estimates*

*Table S 4. Summary of fixed and random effects parameter estimates for MAIHDA mixed-effects regression models presented in main text Figure 3, estimating associations between health & social care access issues and combinations of sex, education level and local area deprivation.*

| | **Model A - Outcome: Number of health & social care access issues (linear regression)** | | | **Model B - Outcome: One or more health & social care access issues (logistic regression)** | | |
|---|---|---|---|---|---|---|
| **Fixed effects: Regression coefficients** | **Estimate** | **95% CI** | **p** | **Odds Ratio** | **95% CI** | **p** |
| (Intercept) | 0.49 | 0.36 – 0.61 | <0.001 | 0.37 | 0.28 – 0.49 | <0.001 |
| Age group [18-39] | 0 | -0.12 – 0.12 | 0.982 | 0.79 | 0.58 – 1.07 | 0.121 |
| Age group [40-49] | -0.08 | -0.17 – 0.00 | 0.063 | 0.91 | 0.73 – 1.13 | 0.394 |
| Age group [50-59] (Reference) | 0 | | | 1 | | |
| Age group [60-69] | -0.14 | -0.21 – -0.07 | <0.001 | 0.78 | 0.65 – 0.94 | 0.01 |
| Age group [≥70] | -0.08 | -0.19 – 0.03 | 0.138 | 0.87 | 0.66 – 1.13 | 0.3 |
| Sex [Female] (Reference) | 0 | | | 1 | | |
| Sex [Male] | -0.09 | -0.19 – 0.01 | 0.09 | 0.83 | 0.65 – 1.06 | 0.13 |
| Ethnic group [Any other ethnic group] | -0.08 | -0.34 – 0.17 | 0.513 | 0.97 | 0.51 – 1.84 | 0.931 |
| Ethnic group [Asian or Asian British] | -0.01 | -0.32 – 0.31 | 0.957 | 0.91 | 0.41 – 2.01 | 0.812 |
| Ethnic group [Black/Black British] | -0.28 | -0.75 – 0.18 | 0.237 | 0.37 | 0.08 – 1.64 | 0.189 |
| Ethnic group [Mixed/multiple] | -0.12 | -0.41 – 0.17 | 0.422 | 0.81 | 0.38 – 1.72 | 0.582 |
| Ethnic group [White] (reference) | 0 | | | 1 | | |
| Highest educational qualification [Prefer not to answer/not stated] | 0.42 | 0.15 – 0.68 | 0.002 | 1.88 | 1.02 – 3.47 | 0.043 |
| Highest educational qualification [Less than University degree or equivalent] | 0.03 | -0.09 – 0.15 | 0.598 | 1.26 | 0.96 – 1.64 | 0.091 |
| Highest educational qualification [University degree] (reference) | 0 | | | 1 | | |
| Highest educational qualification [Postgraduate degree or higher] | -0.07 | -0.18 – 0.05 | 0.246 | 0.89 | 0.68 – 1.16 | 0.38 |
| Local area deprivation [IMD Quintile 1] | 0.17 | -0.00 – 0.35 | 0.051 | 1.69 | 1.14 – 2.50 | 0.008 |
| Local area deprivation [IMD Quintile 2] | 0.14 | -0.00 – 0.29 | 0.056 | 1.53 | 1.09 – 2.14 | 0.013 |
| Local area deprivation [IMD Quintile 3] | 0.07 | -0.07 – 0.21 | 0.325 | 1.23 | 0.90 – 1.68 | 0.194 |
| Local area deprivation [IMD Quintile 4] | 0.09 | -0.05 – 0.22 | 0.195 | 1.22 | 0.89 – 1.67 | 0.216 |
| Local area deprivation [IMD Quintile 5] (reference) | 0 | | | 1 | | |
| **Random effects: Variances** | | | | | | |
| Stratum level | 0.0077 | | | 0.032 | | |
| Individual level | 0.80 | | | 3.29 | | |
| **Summary statistics** | | | | | | |
| Variance partition coefficient | 0.958% | | | 0.956% | | |
| N, strata | 30 | | | 30 | | |
| N, individuals | 3516 | | | 3516 | | |
| Marginal $R^2$ / Conditional $R^2$ | 0.016 / 0.026 | | | 0.022 / 0.032 | | |

### Supplementary MAIHDA intersectional models

*Sample stratified by COVID-19 history*

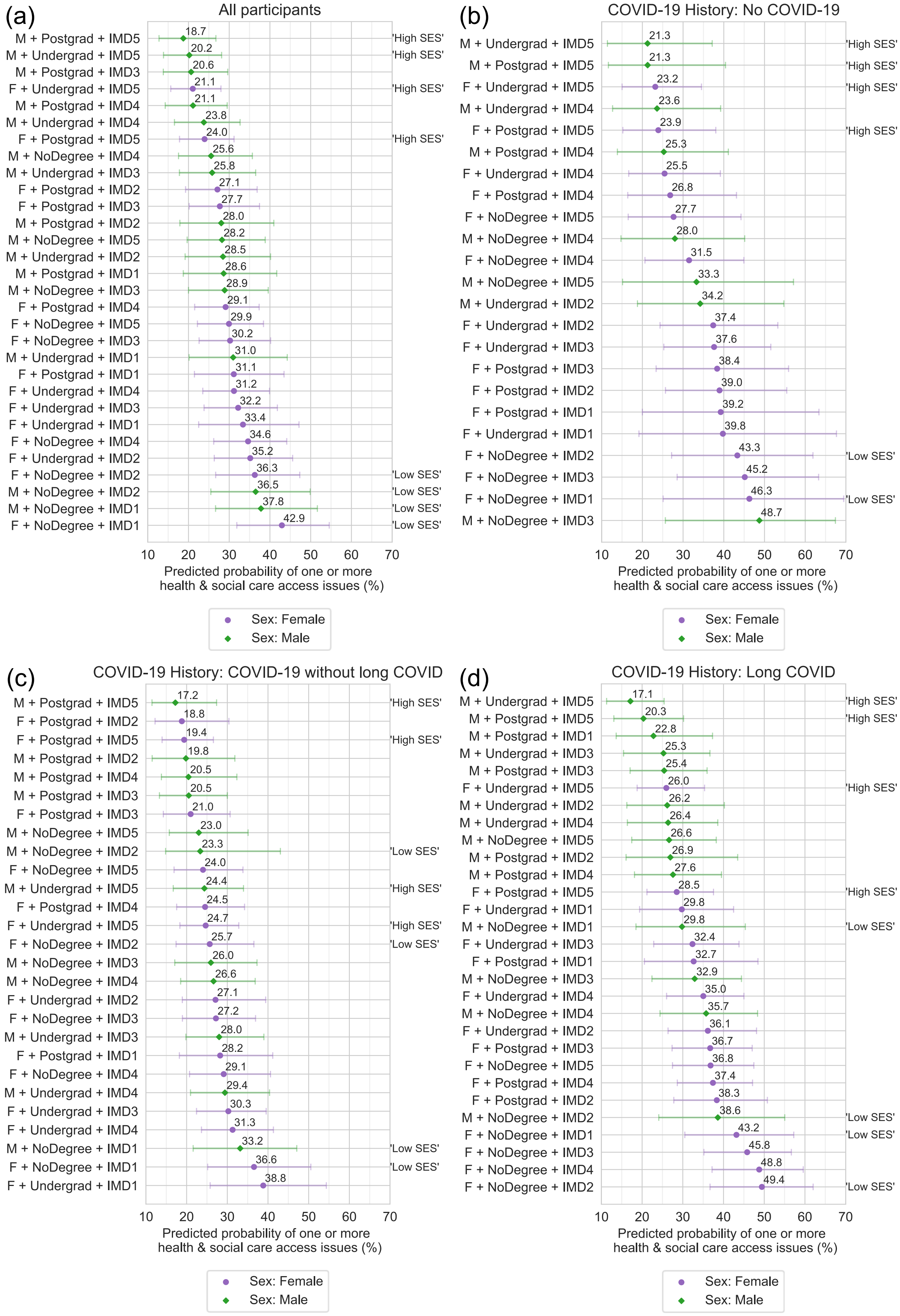

*Figure S 3. Intersectional MAIHDA models estimating effect of combinations of sex, education level and local area deprivation on health & social care access issues during the COVID-19 pandemic, after stratification by COVID-19 history. Predicted probability (with approximate 95% confidence intervals) of one or more issues from logistic regression for: (a) all questionnaire participants (N = 3,516) (as presented in main text Figure 3 (b)), (b) participants with no history of COVID-19 (N = 598), (c) participants with COVID-19 history but no self-reported or diagnosed long COVID (N = 1,647), (d) participants with self-reported or diagnosed long COVID (N = 1,121). 95% confidence intervals are approximate only because the model assumes no sampling covariability between the regression coefficients and stratum random effects. Fixed effects included in models were age group, ethnic group, sex, education level and local area deprivation. Models included participant weighting for inverse probability of questionnaire completion and COVID-19 history. Strata labels: F = Female, M = Male; NoDegree = Less than degree level education (including not stated/prefer not to say), Undergrad = Undergraduate degree level, Postgrad = Postgraduate degree level or higher; IMD1-IMD5 = Index of Multiple Deprivation Quintile 1 to 5, where 1 is most deprived 20% of areas, and 5 is least deprived 20%. Labels "High SES" are given to highlight strata where the combination of education level and deprivation suggest relatively high socio-economic status - living in the least deprived quintile 1 and having a degree level education or higher. "Low SES" highlights relatively low socio-economic status strata – living in the most deprived 40% (quintile 4 or 5) and having less than degree level education. Results from strata with either less than 5 participants or no participants reporting the outcome are not presented in figures.*

*With income included in strata*

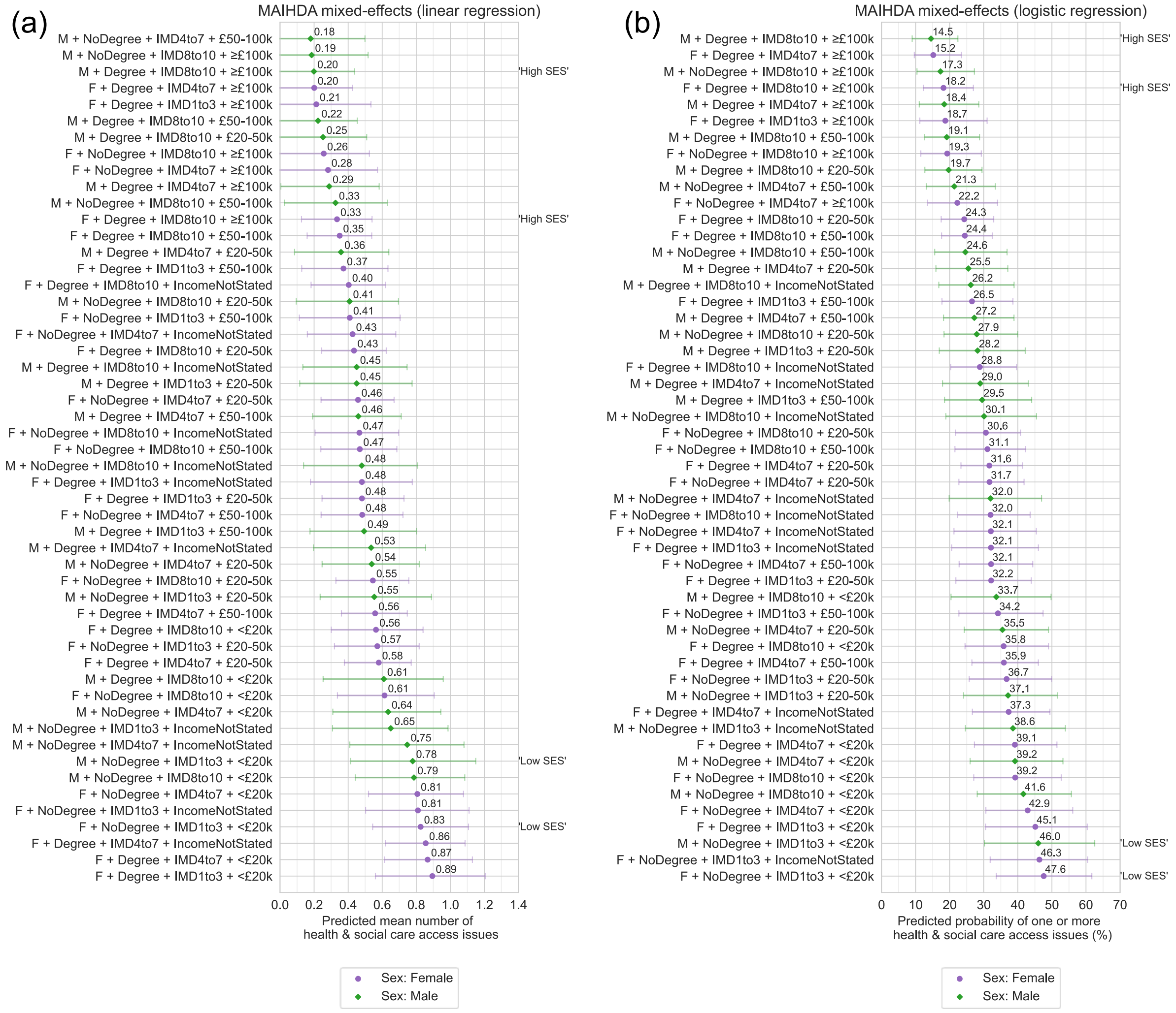

*Figure S 4. Intersectional MAIHDA models estimating effect of combinations of sex, education level, local area deprivation and household income on health & social care access issues during the COVID-19 pandemic. (a) Predicted mean (with approximate 95% confidence intervals) number of issues from linear regression and (b) predicted probability (with approximate 95% confidence intervals) of one or more issues from logistic regression. 95% confidence intervals are approximate only because the model assumes no sampling covariability between the regression coefficients and stratum random effects. Fixed effects included in models were age group, ethnic group, sex, education level, local area deprivation and household income. Models included participant weighting for inverse probability of questionnaire completion. Strata labels: F = Female, M = Male; NoDegree = Less than degree level education (including not stated/prefer not to say), Degree = Undergraduate degree level or higher; IMD1to3 = Index of Multiple Deprivation (IMD) Decile 1 to 3, IMD4to7 = IMD Decile 4 to 7, IMD8to10 = IMD Decile 8 to 10, where 1 is most deprived 10% of areas, and 10 is least deprived 10%; IncomeNotStated = Income question not answered or 'prefer not to say' selected. Labels "High SES" are given to highlight strata where the combination of education level, deprivation and income suggest relatively high socio-economic status - living in the least deprived 30%, having a degree level education or higher, and a household income of £100,000 or more. "Low SES" highlights relatively low socio-economic status strata – living in the most deprived 30% and having less than degree level education and household income of less than £20,000. Results from strata with either less than 5 participants or no participants reporting the outcome are not presented in figures (8 of 60 strata).*

#### *With pre-pandemic health factors as additional adjustment variables*

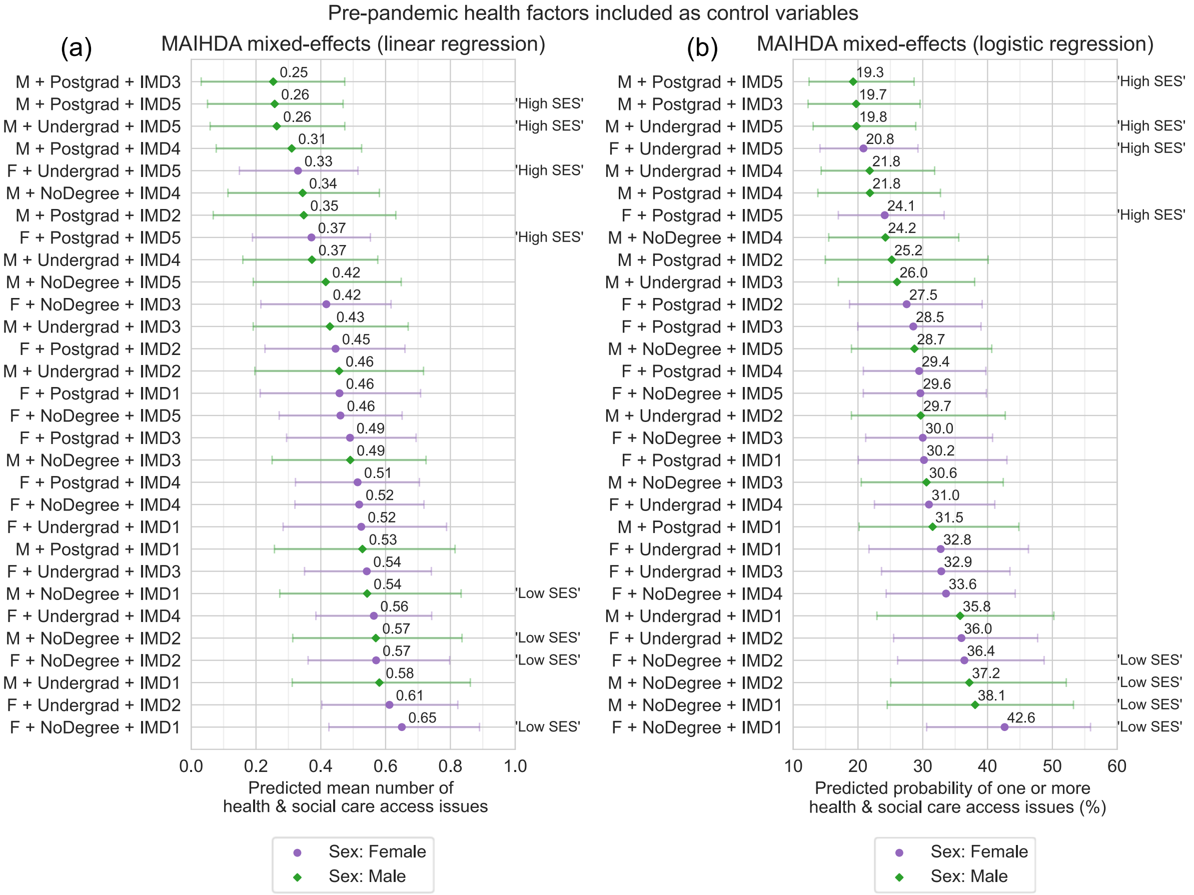

*Figure S 5. Intersectional MAIHDA models estimating effect of combinations of sex, education level and local area deprivation on health & social care access issues during the COVID-19 pandemic, including adjustment for pre-pandemic health factors. (a) Predicted mean (with approximate 95% confidence intervals) number of issues from linear regression and (b) predicted probability (with approximate 95% confidence intervals) of one or more issues from logistic regression. Fixed-effects included in models were age group, ethnic group, sex, education level, local area deprivation, pre-pandemic general health, body mass index, PRISMA-7 frailty score, and number of long-term physical health conditions. Models included participant weighting for inverse probability of questionnaire completion. Strata labels: F = Female, M = Male; NoDegree = Less than degree level education (including not stated/prefer not to say), Undergrad = Undergraduate degree level, Postgrad = Postgraduate degree level or higher; IMD1-IMD5 = Index of Multiple Deprivation Quintile 1 to 5, where 1 is most deprived 20% of areas, and 5 is least deprived 20%. Labels "High SES" are given to highlight strata where the combination of education level and deprivation suggest relatively high socio-economic status - living in the least deprived quintile 1 and having a degree level education or higher. "Low SES" highlights relatively low socio-economic status strata – living in the most deprived 40% (quintile 4 or 5) and having less than degree level education.*

#### Qualitatively coded subset full codes & frequencies

*Table S 5. Unadjusted frequencies of qualitative codes used to categorise open-text responses about COVID-19 health care experiences in part 2 of the study, split by socio-demographic group. Codes with frequency of less than 5 are masked.*

| | | | | | Relatively disadvantaged | | Intermediate | | Relatively advantaged | | Total | |
|---|---|---|---|---|---|---|---|---|---|---|---|---|
| Code | Topic | Level 1 | Level 2 | Level 3 | N | % | N | % | N | % | N | % |
| | | | | **Total sample size** | **109** | **100%** | **117** | **100%** | **109** | **100%** | **335** | **100%** |
| 1 | Reflection on health care experiences: | Positive experience | | | 16 | 15% | 17 | 15% | 12 | 11% | 45 | 13% |
| 2 | | | But it didn't help recovery from COVID illness | | <5 | - | <5 | - | <5 | - | <5 | - |
| 3 | | | And it did help recovery from illness | | 5 | 5% | 5 | 4% | 6 | 6% | 16 | 5% |
| 4 | | Neutral/mixed | | | 8 | 7% | 11 | 9% | 14 | 13% | 33 | 10% |
| 5 | | Unclear/not possible to discern | | | 25 | 23% | 13 | 11% | 16 | 15% | 54 | 16% |
| 6 | | Negative experience | | | 29 | 27% | 25 | 21% | 12 | 11% | 66 | 20% |
| 7 | | | But it didn't impact recovery from illness negatively | | <5 | - | <5 | - | <5 | - | <5 | - |
| 8 | | | Sense it did impact recovery from illness negatively | | 6 | 6% | 5 | 4% | <5 | - | 15 | 4% |
| 9 | Description of experiences: | Did not receive care for COVID-19 | | | 50 | 46% | 61 | 52% | 52 | 48% | 163 | 49% |
| 10 | | | Did not attempt to contact health care services… | | 24 | 22% | 37 | 32% | 46 | 42% | 107 | 32% |
| 11 | | | | but wanted or needed to | <5 | - | <5 | - | <5 | - | 8 | 2% |
| 12 | | | | and did not need them | 13 | 12% | 20 | 17% | 30 | 28% | 63 | 19% |
| 13 | | | Attempted to use health care services but did not receive care, because: | | 11 | 10% | 5 | 4% | <5 | - | 20 | 6% |
| 14 | | | | Unable to get appointment/into primary care/GP | 11 | 10% | 5 | 4% | <5 | - | 20 | 6% |
| 15 | | | | Not able to attend appointment | <5 | - | <5 | - | <5 | - | <5 | - |
| 16 | | Received care for COVID-19 | | | 50 | 46% | 50 | 43% | 43 | 39% | 143 | 43% |
| 17 | | | Received primary care/GP | | 41 | 38% | 34 | 29% | 35 | 32% | 110 | 33% |
| 18 | | | Received secondary care/hospital | | 19 | 17% | 21 | 18% | 14 | 13% | 54 | 16% |
| 19 | | | Received care in 'long covid clinic' or from specialist | | <5 | - | 6 | 5% | 8 | 7% | 18 | 5% |

| | | | | | | | | | | | | |
|---|---|---|---|---|---|---|---|---|---|---|---|---|
| 20 | | | Received social/community care | | <5 | - | <5 | - | <5 | - | <5 | - |
| 21 | | | Accessing was easy/straightforward (explicit or inferred) | | 12 | 11% | 8 | 7% | 16 | 15% | 36 | 11% |
| 22 | | | Accessing was difficult (explicit or inferred) | | 10 | 9% | 12 | 10% | 10 | 9% | 32 | 10% |
| 23 | | | Used private health care | | <5 | - | <5 | - | 6 | 6% | 12 | 4% |
| 24 | | | Received alternative care (ex. homeopathy) | | <5 | - | <5 | - | <5 | - | <5 | - |
| 25 | | | Received care (perceived as) inadequate - care received was unhelpful or irrelevant, had appointment but effectively no care provided, etc. | | 16 | 15% | 10 | 9% | 8 | 7% | 34 | 10% |
| 27 | | Mention of influences that related to or affected experiences with care (whether or not care was received) | Personal time constraints – e.g. not allowed time off work, caring/other responsibilities | | <5 | - | <5 | - | <5 | - | <5 | - |
| 28 | | | Financial constraints – e.g. couldn’t afford time off work, private care etc | | <5 | - | <5 | - | <5 | - | <5 | - |
| 29 | | | Benefitted from resources – e.g. benefitted from being able to afford private care, had time to try multiple times to access care | | <5 | - | 6 | 5% | 7 | 6% | 16 | 5% |
| 30 | | | ‘Inside knowledge’ of health care system – e.g. personal medical knowledge or NHS experience, or family/friends with experience in NHS, helping to navigate system & communicate with health care professionals | | <5 | - | 7 | 6% | 9 | 8% | 18 | 5% |
| 31 | | | Stigma & Discrimination experiences – sense that person experienced stigma or discrimination in health care settings because of who they were | | <5 | - | <5 | - | <5 | - | <5 | - |
| 32 | | | Negative interpersonal experiences - patient feels disbelieved, treated rudely, 'gaslit', etc. | | 5 | 5% | 8 | 7% | <5 | - | 17 | 5% |
| 33 | | | Absence/lack of trust in health care services - patients' pre-existing assumptions about the | | <5 | - | <5 | - | <5 | - | 8 | 2% |

| | | | |
|---|---|---|---|
| | | | HC system influencing their access behaviours |

#### Qualitatively coded subset socio-demographic characteristics

*Table S 6. Socio-demographic characteristics of qualitatively coded subset of participants, split by socio-demographic group. Figures for group sizes of less than 5 are suppressed.*

| | | Relatively disadvantaged | | Intermediate | | Relatively advantaged | | Total | |
|---|---|---|---|---|---|---|---|---|---|
| Domain | Variable | N | % | N | % | N | % | N | % |
| | **Total** | 109 | | 117 | | 109 | | 335 | |
| Individual pre-pandemic demographics | **Age (median, interquartile range)** | 58 (52-64) | | 58 (51-66) | | 58 (51-63) | | 58 (51-65) | |
| | **Age group (years)** | | | | | | | | |
| | 18-39 | <5 | - | 9 | 7.7% | <5 | - | 14 | 4.2% |
| | 40-49 | 16 | 14.7% | 18 | 15.4% | 19 | 17.4% | 53 | 15.8% |
| | 50-59 (reference) | 49 | 45.0% | 30 | 25.6% | 43 | 39.4% | 122 | 36.4% |
| | 60-69 | 31 | 28.4% | 42 | 35.9% | 33 | 30.3% | 106 | 31.6% |
| | ≥ 70 | 12 | 11.0% | 18 | 15.4% | 10 | 9.2% | 40 | 11.9% |
| | **Sex** | | | | | | | | |
| | Female (reference) | 93 | 85.3% | 98 | 83.8% | 87 | 79.8% | 278 | 83.0% |
| | Male | 16 | 14.7% | 19 | 16.2% | 22 | 20.2% | 57 | 17.0% |
| | **Ethnic group** | | | | | | | | |
| | Asian/Asian British | 5 | 4.6% | <5 | - | <5 | - | 6 | 1.8% |
| | Black/Black British | <5 | - | <5 | - | <5 | - | <5 | - |
| | Mixed/Multiple | 5 | 4.6% | <5 | - | <5 | - | 5 | 1.5% |
| | Other | 11 | 10.1% | <5 | - | <5 | - | 13 | 3.9% |
| | White (reference) | 84 | 77.1% | 114 | 97.4% | 109 | 100.0% | 307 | 91.6% |
| | **First language** | | | | | | | | |
| | English (reference) | 104 | 95.4% | 114 | 97.4% | 107 | 98.2% | 325 | 97.0% |
| | Other | <5 | - | <5 | - | <5 | - | 6 | 1.8% |
| | Prefer not to answer/not stated | <5 | - | <5 | - | <5 | - | <5 | - |
| | **Highest educational qualification** | | | | | | | | |
| | Prefer not to answer/not stated | <5 | - | <5 | - | <5 | - | 6 | 1.8% |
| | Did not complete secondary school | <5 | - | <5 | - | <5 | - | 5 | 1.5% |
| | GCSE or GNVQ or equivalent | 43 | 39.4% | 22 | 18.8% | <5 | - | 65 | 19.4% |
| | A-Levels or advanced GNVQ or equivalent | 46 | 42.2% | 25 | 21.4% | <5 | - | 71 | 21.2% |
| | University degree (reference) | 11 | 10.1% | 51 | 43.6% | <5 | - | 65 | 19.4% |
| | Postgraduate degree or higher | <5 | - | 15 | 12.8% | 94 | 86.2% | 111 | 33.1% |
| | PhD | <5 | - | <5 | - | 12 | 11.0% | 12 | 3.6% |
| | **UK Region** | | | | | | | | |
| | East Midlands | 10 | 9.2% | <5 | - | <5 | - | 18 | 5.4% |
| | East of England | 11 | 10.1% | 13 | 11.1% | 15 | 13.8% | 39 | 11.6% |
| | London (reference) | 27 | 24.8% | 28 | 23.9% | 21 | 19.3% | 76 | 22.7% |
| | North East | <5 | - | <5 | - | <5 | - | 7 | 2.1% |
| | North West | 14 | 12.8% | 7 | 6.0% | 8 | 7.3% | 29 | 8.7% |
| | Scotland & Northern Ireland | 5 | 4.6% | <5 | - | <5 | - | 9 | 2.7% |
| | South East | 10 | 9.2% | 24 | 20.5% | 29 | 26.6% | 63 | 18.8% |
| | South West | 6 | 5.5% | 16 | 13.7% | 10 | 9.2% | 32 | 9.6% |
| | Wales | 5 | 4.6% | 6 | 5.1% | <5 | - | 15 | 4.5% |
| | West Midlands | 9 | 8.3% | 6 | 5.1% | 8 | 7.3% | 23 | 6.9% |

| | | | | | | | | | |
|---|---|---|---|---|---|---|---|---|---|
| | Yorkshire and The Humber | 8 | 7.3% | 10 | 8.5% | 6 | 5.5% | 24 | 7.2% |
| | **Pre-pandemic employment status** | | | | | | | | |
| | Employed (reference) | 63 | 57.8% | 66 | 56.4% | 57 | 52.3% | 186 | 55.5% |
| | Self-employed | 10 | 9.2% | 11 | 9.4% | 21 | 19.3% | 42 | 12.5% |
| | Unemployed | <5 | - | <5 | - | <5 | - | <5 | - |
| | Permanently or long-term sick or disabled | 12 | 11.0% | <5 | - | <5 | - | 14 | 4.2% |
| | Retired | 14 | 12.8% | 29 | 24.8% | 18 | 16.5% | 61 | 18.2% |
| | Other | 7 | 6.4% | 7 | 6.0% | 11 | 10.1% | 25 | 7.5% |
| | Unknown | <5 | - | <5 | - | <5 | - | 6 | 1.8% |
| | **Rural-Urban classification** | | | | | | | | |
| | Rural | 9 | 8.3% | 32 | 27.4% | 30 | 27.5% | 71 | 21.2% |
| | Urban (reference) | 100 | 91.7% | 85 | 72.6% | 79 | 72.5% | 264 | 78.8% |
| | **Local area deprivation** | | | | | | | | |
| | IMD Decile 1 (most deprived 10%) | 24 | 22.0% | <5 | - | <5 | - | 26 | 7.8% |
| | IMD Decile 2 | 26 | 23.9% | <5 | - | <5 | - | 30 | 9.0% |
| | IMD Decile 3 | 32 | 29.4% | 9 | 7.7% | <5 | - | 41 | 12.2% |
| | IMD Decile 4 | 7 | 6.4% | 10 | 8.5% | <5 | - | 19 | 5.7% |
| | IMD Decile 5 | <5 | - | 14 | 12.0% | <5 | - | 18 | 5.4% |
| | IMD Decile 6 | <5 | - | 11 | 9.4% | <5 | - | 14 | 4.2% |
| | IMD Decile 7 | <5 | - | 19 | 16.2% | <5 | - | 25 | 7.5% |
| | IMD Decile 8 | <5 | - | 16 | 13.7% | 30 | 27.5% | 49 | 14.6% |
| | IMD Decile 9 | <5 | - | 14 | 12.0% | 39 | 35.8% | 57 | 17.0% |
| | IMD Decile 10 (least deprived 10%) (reference) | <5 | - | 18 | 15.4% | 35 | 32.1% | 56 | 16.7% |
| **Socio-demographics during pandemic** | **Current household income** | | | | | | | | |
| | Prefer not to answer/not stated | 21 | 19.3% | 21 | 17.9% | <5 | - | 43 | 12.8% |
| | Less than £20,000 | 28 | 25.7% | 8 | 6.8% | <5 | - | 36 | 10.7% |
| | £20,000-£29,999 | 15 | 13.8% | 13 | 11.1% | <5 | - | 30 | 9.0% |
| | £30,000-£39,999 | 14 | 12.8% | 13 | 11.1% | <5 | - | 27 | 8.1% |
| | £40,000-£49,999 | 12 | 11.0% | 11 | 9.4% | <5 | - | 26 | 7.8% |
| | £50,000-£74,999 (reference) | 10 | 9.2% | 28 | 23.9% | <5 | - | 42 | 12.5% |
| | £75,000-£99,999 | 9 | 8.3% | 16 | 13.7% | 5 | 4.6% | 30 | 9.0% |
| | £100,000 or more | <5 | - | 7 | 6.0% | 94 | 86.2% | 101 | 30.1% |

Qualitatively coded subset poisson regression model results

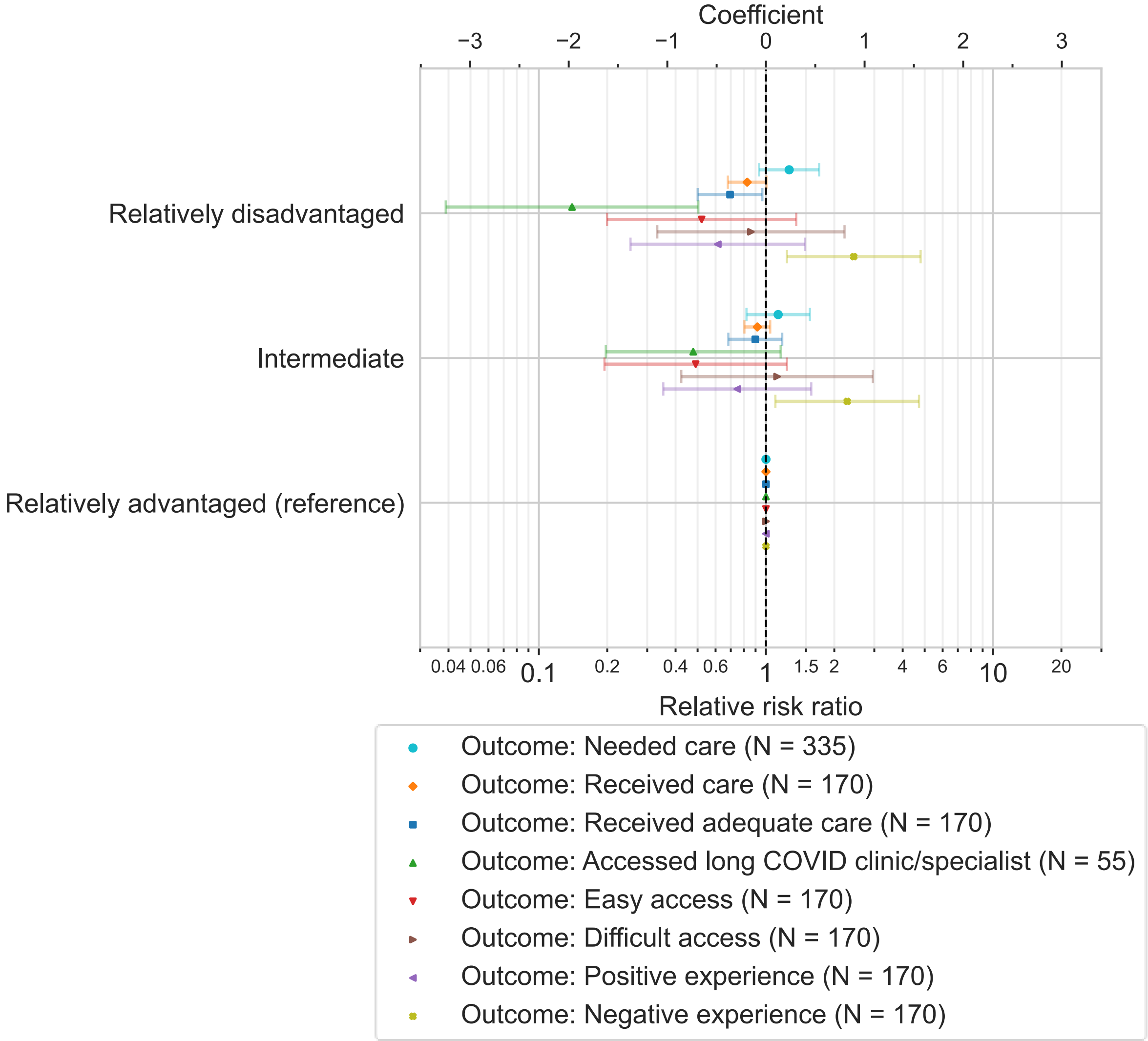

*Figure S 6. Results of multivariable poisson regression estimating the likelihood of various health experience outcomes for socio-demographic groups within the qualitatively coded subset. Models adjusted for respondent age group, and weighted for inverse probability of selection into samples. The sample size of the subset used to model each outcome are given in the legend.*

References

1 List of ethnic groups - GOV.UK. https://www.ethnicity-facts-figures.service.gov.uk/style-guide/ethnic-groups (accessed 2 Mar 2023).

2 English indices of deprivation 2019 - GOV.UK. https://www.gov.uk/government/statistics/english-indices-of-deprivation-2019 (accessed 29 Nov 2021).

3 Welsh Index of Multiple Deprivation (full Index update with ranks): 2019 | GOV.WALES. https://gov.wales/welsh-index-multiple-deprivation-full-index-update-ranks-2019 (accessed 29 Nov 2021).

4 Scottish Index of Multiple Deprivation 2020 - gov.scot. https://www.gov.scot/collections/scottish-index-of-multiple-deprivation-2020/ (accessed 29 Nov 2021).

5 Northern Ireland Multiple Deprivation Measure 2017 (NIMDM2017) | Northern Ireland Statistics and Research Agency. https://www.nisra.gov.uk/statistics/deprivation/northern-ireland-multiple-deprivation-measure-2017-nimdm2017 (accessed 29 Nov 2021).

6 Xue Q-L. The Frailty Syndrome: Definition and Natural History. *Clinics in Geriatric Medicine* 2011;**27**:1–15. doi:10.1016/j.cger.2010.08.009

7 Raîche M, Hébert R, Dubois MF. PRISMA-7: A case-finding tool to identify older adults with moderate to severe disabilities. *Archives of Gerontology and Geriatrics* 2008;**47**:9–18. doi:10.1016/J.ARCHGER.2007.06.004
